# Single-cell profiling of BAL in preschool cystic fibrosis reveals macrophage dysregulation and ivacaftor-modified inflammatory programs in the early life lung

**DOI:** 10.1101/2025.02.24.25322508

**Authors:** Jovana Maksimovic, Shivanthan Shanthikumar, George Howitt, Gunjan Dixit, Peter F Hickey, Casey Anttila, Daniel V. Brown, Liam Gubbels, Anne Senabouth, Daniela Amann-Zalcenstein, Joseph E. Powell, Sarath C. Ranganathan, Alicia Oshlack, Melanie R. Neeland

## Abstract

Aberrant inflammation and structural lung damage occurs early in life for people with cystic fibrosis (CF). Even in the era of CFTR modulators, anti-inflammatory therapy may still be needed to prevent establishment and lifelong consequences of bronchiectasis. In this study, we integrated transcriptome-wide single-cell RNA sequencing data and highly multiplexed surface protein expression to create the largest comprehensive paediatric lower airway atlas of >190,000 cells from 45 bronchoalveolar lavage (BAL) samples resulting in 43 immune and epithelial cell populations. We then investigated inflammatory cell responses in children with CF to show widespread gene expression dysregulation of macrophage populations in the preschool CF lung. This included alterations in pathways associated with TNF and IFN signalling, cholesterol homeostasis, as well as pulmonary fibrosis, that were further altered by the early development of bronchiectasis. We showed that the CFTR modulator ivacaftor restores some of these macrophage-related functional deficits and reduces expression of pathways associated with neutrophil infiltration, however the modulator lumacaftor/ivacaftor did not result in any detectable changes in transcriptional response. This work represents a comprehensive, multi-omic single-cell analysis of bronchoalveolar lavage from preschool children and the results inform the future development of anti-inflammatory therapy for children with CF.

## INTRODUCTION

Cystic fibrosis (CF) is an autosomal recessive life-shortening condition, caused by mutations in the cystic fibrosis transmembrane conductance regulator (*CFTR*) gene ^1^. While it is a multisystem disease, lung disease which specifically affects the airways (the site of *CFTR* expression) ^2^, is the major source of morbidity and mortality. Lung disease results from an interplay of infection and inflammation which begin in early life ^3^. As the disease progresses, infections can become chronic, and neutrophil dominated inflammation occurs. However, knowledge gaps remain regarding the early stages of CF lung disease and the defective mechanisms that result in chronic infection and neutrophilic inflammation.

Single cell RNA sequencing (sc-RNAseq) has allowed investigation of biological processes in both health and disease, at a resolution which was previously not possible. The application of sc-RNAseq to the investigation of CF lung disease has yielded several findings, including the exact distribution of *CFTR* expression in the airway ^2^, altered airway epithelial cell type proportions ^4^, increased abundance of immature and proinflammatory neutrophils ^5^, increased activation of adaptive immune cells ^6^, unique communication between epithelial, immune and endothelial cells ^6^, and the ability of CFTR directed therapies to restore defects in epithelial and immune cell function ^7^.

As emphasised in a recent review by Januska and Walsh ^8^, which underscored the importance of including infants and young children in future investigations, scRNA-seq studies to date have largely focused on samples from adults with advanced lung disease, where epithelial cells predominate. As such, sc-RNAseq has not been applied to assess the earliest stages of CF lung disease and the role of immune cells in the establishment of chronic infection and inflammation. It is important to understand the earliest manifestations of the disease because irreversible structural lung disease develops in this period ^9^. Once that has occurred, even if CFTR dysfunction is corrected, via either CFTR modulators or novel approaches such as gene therapy, people with CF will still have to manage the lifelong consequences of bronchiectasis ^10^.

In particular, sc-RNAseq has not been used to robustly evaluate the role of alveolar macrophages (AMs) in early stages of CF lung disease. AMs are the predominant immune cell in the lung and have several crucial functions, including phagocytosis of debris, pathogen detection, regulation of the innate immune response, leukocyte recruitment and resolution of inflammation ^11^. The AM pool is made up of self-replenishing tissue resident AM (derived from yolk sac progenitors), and bone marrow derived AMs which are recruited during inflammatory responses (either acute or chronic). AMs are a functionally heterogenous population influenced by factors such as origin (tissue resident vs, bone marrow derived) and pathogen exposure. However, until recently it has been difficult to evaluate this heterogeneity and prior attempts at classifying macrophage heterogeneity were significantly flawed ^12^. sc-RNA-seq has been able to unravel AM heterogeneity in health ^13^ and lung diseases such as pneumonia ^14^, chronic obstructive pulmonary disease ^15^, and granulomatous lung disease ^16^, but not yet in early CF. Given AMs regulate immune defence and airway inflammation, they are likely to play a key role in the early pathogenesis of CF lung disease especially prior to the onset to neutrophil dominated inflammation. Prior studies using animal models, or monocytes as a surrogate of AMs, have shown several defects including impaired phagocytosis and bacterial killing and altered inflammatory response ^17^ . However, there are significant challenges with the models used to date ^18^ and a clear need to investigate AMs from people with CF.

In this study, we profiled over 190,000 high-quality cells from 45 bronchoalveolar lavage (BAL) samples collected from preschool children with cystic fibrosis (CF) and age-matched non-CF controls, generating the largest single-cell dataset of the paediatric airway to date. Relative to previously investigated samples such as blood (which profiles the systemic immune environment) and airway brushings (which are dominated by epithelial cells), BAL is the ideal sample to assess lung inflammation and resident immune cells such as AMs. The macrophage-rich composition of BAL samples allowed us to comprehensively examine this population in preschool children with CF. The CF cohort included children both receiving and not receiving CFTR modulator therapy, as well as those with and without radiographic evidence of bronchiectasis. We applied best-practice pseudobulk differential expression analysis ^19^ to control for inter-individual variability and confounding factors. This enabled the identification of gene expression changes associated with CF disease status, severity, and treatment. Our findings provide new insights into immune cell contributions during the early stages of CF lung disease and offer a framework to guide the development of targeted anti-inflammatory therapies.

## RESULTS

### 1. An immune and epithelial cell atlas of early life BAL

This study includes sc-RNAseq analysis of 45 BAL samples from 28 children with CF and 8 non-CF controls (6 of the children with CF were sampled multiple times, see Figure 1A and Extended Data File 1). The CF group consisted of children who had surveillance bronchoscopy as part of clinical care [5], and the control group were children who underwent general anaesthesia for assessment of upper airway disease (i.e. stridor). The median age of the CF participants was 3.03 years (range 0.5-6.2 years) and the median age of the non-CF controls was 2.11 years (range 0.84-8.4 years). For children with CF, metadata were recorded including treatment with modulator therapy (ivacaftor or lumacaftor/ivacaftor), pathogen detection in BAL, and presence or absence of bronchiectasis. Single-cell RNA sequencing of BAL samples was performed using the 3’ 10x Genomics Chromium platform, either with or without the addition of ADTs (BioLegend TotalSeqA Human Universal Cocktail v1.0). Details of the bioinformatic analysis undertaken are provided in the methods and supplement, and the complete analysis is available at https://oshlacklab.com/paediatric-cf-inflammation-citeseq/index.html with additional data tables available at https://github.com/Oshlack/paediatric-cf-inflammation-citeseq/tree/main/output/dge_analysis.

**Figure 1.**
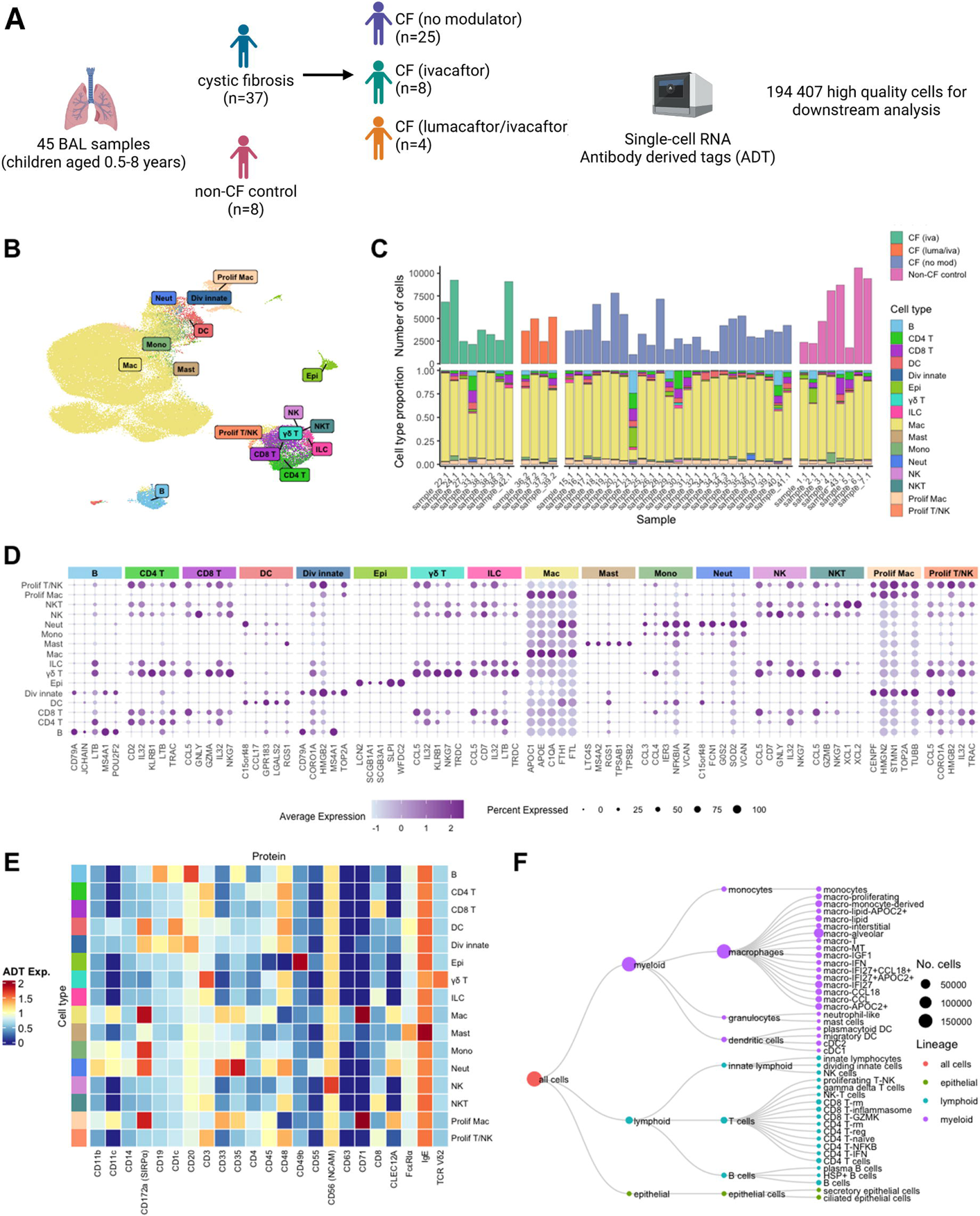
An immune and epithelial cell atlas of early life BAL. (A) Overview of study cohort and experimental workflow. (B) UMAP depicting the broad immune and epithelial cell populations identified in early life BAL; based on dimension reduction of gene expression data only. (C) Bar plots depicting total cell numbers for each BAL sample (top), and the proportion of each broad cell type within each BAL sample (bottom), separated by treatment and disease groups. (D) Dot plot depicting marker genes of each identified broad immune and epithelial cell population. (E) Heatmap depicting protein (antibody derived tag (ADT)) expression on each identified broad immune and epithelial cell population. (F) Hierarchy of cell types identified in this study. The size of the circle at each level indicates the number of cells of that type.

Unsupervised clustering of sc-RNAseq data and cell annotation using marker gene analysis revealed 16 transcriptionally distinct broad cell populations in paediatric BAL representing all the expected cell types (Figure 1B-C). Marker genes for each broad cell population are shown in Figure 1D. These marker genes included those described previously for well characterised lineages ^20,21^. We found that most cells within the BAL samples were macrophages, contributing 165,209 or 88.98%. T, B and natural killer cells were also captured but only contributed 27,351 cells or 14.07% of the total. Only 1,847 epithelial cells were captured, which was only 0.95% of all cells (Figure 1C). We also observed three subsets of proliferating cells (annotated proliferating T/NK, proliferating macrophages and dividing innate cells) which expressed known proliferation genes including *TOP2A, MKI67, STMN1*, and *TUBB*. The annotations of each cell population were further confirmed by analysis of expression of TotalSeq-A ADTs (Figure 1E). Macrophages expressed myeloid lineage proteins CD172α, CD11c, and CD71, as previously reported for alveolar macrophages in adults ^22,23^. Monocytes were distinguished from macrophages based on the pan monocyte marker CD14, as well as the lack of CD71 and CD169. Dendritic cells expressed CD11c, CD172α and CD1c; and mast cells expressed known protein markers FcεR1α, CD63, and IgE (33). Cells annotated as lymphoid lineage by marker genes expressed expected lineage proteins, including B cells (CD19+CD20+), CD4 T cells (CD3+ CD4+), CD8 T cells (CD3+ CD8+), γδ T cells (CD3+TCRVδ2+), NK cells (CD3-CD56+) and NK-T cells (CD3+CD56+). Epithelial cells were positive for tissue resident marker CD49b. These results confirm that, compared with prior CF studies using airway brushings or lung tissue, BAL samples yielded a greater proportion of alveolar macrophages (AMs) for downstream analysis (Figure 1F).

### 2. Sub-clustering analysis reveals heterogeneity within BAL macrophages, T cells, myeloid cells, and other rare cell populations

As macrophages are the most abundant immune cell in BAL, we performed a sub-clustering analysis of our broad macrophage population. This revealed 16 subpopulations with distinct transcriptional signatures (Figure 2A-C). Eight of these have been previously reported in studies from adult lung samples ^20,24^. We also show five subpopulations of macrophages that can be defined based on expression of *IFI27* and *APOC2*, two key genes shown in a recent study to define macrophage populations in healthy BAL ^24^. We identified *IFI27+* macrophages (macro-IFI27), *IFI27+* macrophages with upregulation of *CCL18* (macro-IFI27+CCL18+), macrophages positive for both *IFI27* and *APOC2* (macro-IFI27+APOC2+), *APOC2+* macrophages (macro-APOC2+) as well as a subset of macrophages expressing *APOC2+* as well as lipid transport associated genes (macro-lipid-APOC2+). We identified a subset of macrophages enriched for insulin growth factor 1 (*IGF1*) (macro-IGF1), and another subset of macrophages showing evidence of T cell interaction based on marker genes (*CCL5, IL32, CD2*) and protein expression. It is likely that these macro-T cells represent doublets, however a growing body of evidence using a variety of methods have suggested that biologically meaningful insights into cellular interactions could be gained from these populations ^25–28^. Finally, we show one subset of proliferating macrophages. Protein expression on each of these macrophage populations is shown in Supplementary Figure 2A.

**Figure 2.**
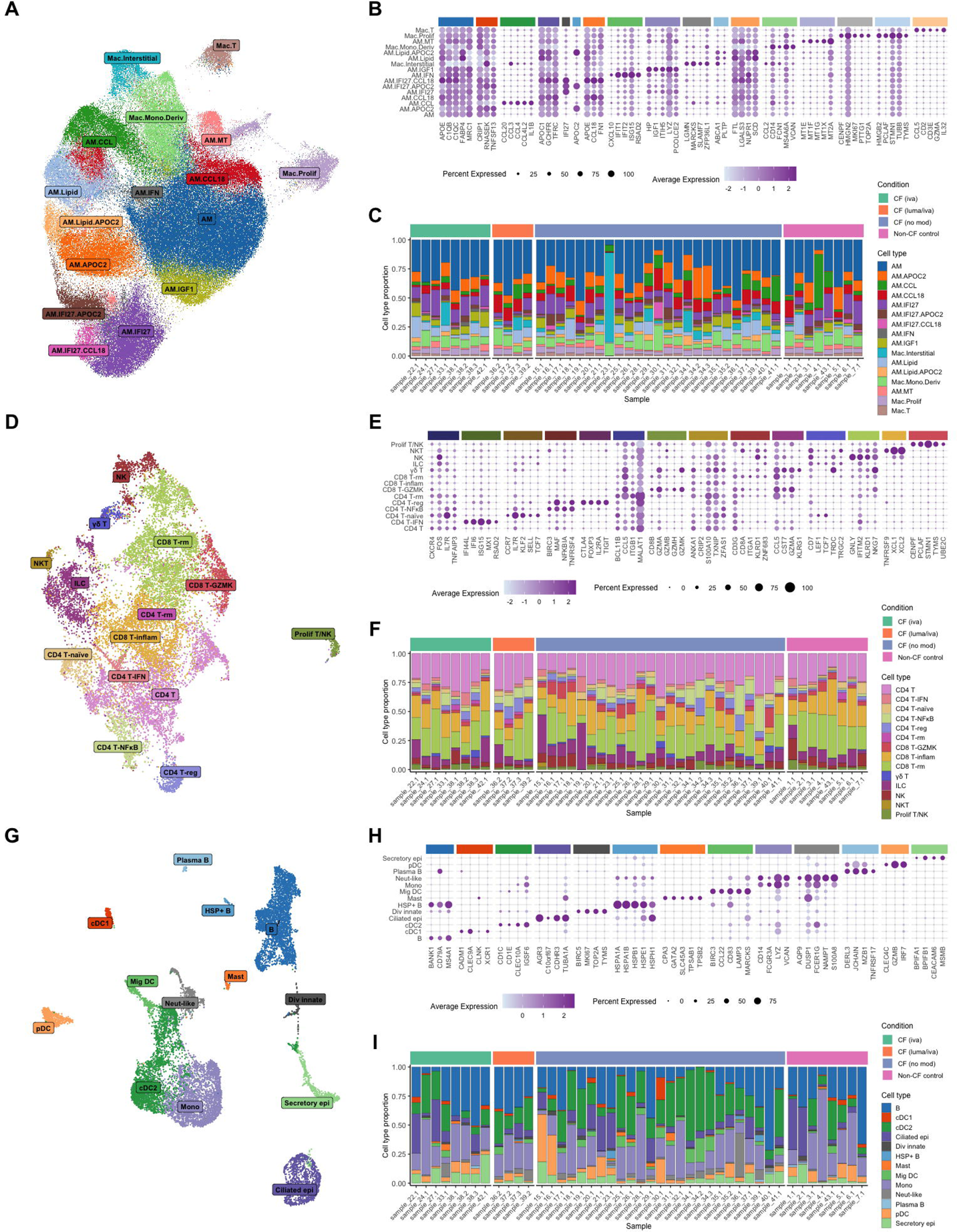
Sub-clustering analysis reveals heterogeneity within BAL macrophages, T cells, myeloid cells, and other rare cell populations. UMAP depicting sub-clustering analysis of BAL macrophages; based on dimension reduction of gene expression data only. (B) Dot plot depicting marker genes of each identified macrophage subpopulation. (C) Stacked bar plot depicting the proportion of each macrophage subpopulation in each BAL sample, separated by treatment and disease groups. (D) UMAP depicting sub-clustering analysis of BAL T/NK cells; based on dimension reduction of gene expression and ADT data. (E) Dot plot depicting marker genes of each identified T/NK cell subpopulation. (F) Stacked bar plot depicting the proportion of each T/NK cell subpopulation in each BAL sample, separated by treatment and disease groups. (G) UMAP depicting sub-clustering analysis of BAL myeloid and rare cell populations; based on dimension reduction of gene expression and ADT data. (H) Dot plot depicting marker genes of each identified myeloid/rare cell subpopulation. (I) Stacked bar plot depicting the proportion of each myeloid/rare subpopulation in each BAL sample, separated by treatment and disease groups.

As with macrophages, we next explored heterogeneity within T/NK subsets which revealed 14 subpopulations with distinct transcriptional signatures, 11 of which have previously been reported (Figure 2D-F) ^20,21^. Two subtypes of CD8 T cells (CD8 Trm and CD8-GZMK) and NK-T cells showed gene and protein markers of lung tissue residency, including surface expression of CD103 and CD49a ^29^ (Figure 2D-F). Conversely, CD4 T cell subsets, NK cells, innate lymphocytes, and γδ T cells were negative or only weakly positive for these markers of tissue residency, suggesting they may have been recently recruited from the circulation. We also show 3 subsets of unique activated T cells, which were annotated based on marker genes as well as REACTOME pathway analysis, one expressing *NF*κ*B* related genes and pathways (CD4 T-NFκB), one expressing *IFN* related genes and pathways (CD4 T-IFN), and one expressing inflammasome activated genes and pathways (CD8 T-inflammasome). To our knowledge, we are the first to describe subtypes of T cells enriched for IFN, NFκB and inflammasome signalling genes in the lung. These inflammatory T cells were observed in both the control and children with CF (Figure 2F). Supplementary Figure 2B shows protein expression across each T/NK cell population, confirming the presence of key lineage markers and providing further validation of our annotations.

A final sub-clustering analysis of all other cells (not macrophages, not T/NK) revealed further heterogeneity within the rarer monocyte, DC, B cell and epithelial cell clusters (Figure 2G-I). DCs were separated into conventional DC (cDC1 and cDC2), plasmacytoid DC, and migratory DC, based on previously described marker genes and protein expression (Supplementary Figure 2C). Plasma B cells were also identified, characterised by key marker genes (*MZB1, TNFRSF17)* as well as protein expression of plasma markers CD27 and CD38. A rare subset of B cells expressing heat shock proteins (HSP) was also identified in our data, characterised by expression of HSP genes *HSPA1B, HSPA6, HSPB1* and enrichment of REACTOME pathways associated with heat shock response. This subset of B cells has recently been described in a single cell sequencing study of rheumatoid arthritis ^30^. We identified 2 subtypes of airway epithelial cells: ciliated epithelial cells and secretory epithelial cells. Protein expression on each of these rare cell populations is shown in Supplementary Figure 2C.

An inherent limitation of our data is that our samples were cryopreserved and thawed prior to analysis. This process is known to deplete granulocytes, which is why we do not see a large population of neutrophils in our dataset, even in the CF samples. As is the case with most clinical studies, our sampling times are opportunistic, and cryopreservation is unavoidable. However, the use of fresh samples would not have obviated issues with identifying neutrophil populations in sc-RNAseq data, as evidenced by the absence of neutrophils in a prior study of CF BAL which used fresh samples ^24^. Nevertheless, we identified a small population of neutrophils expressing key inflammatory genes including S100A8, DUSP1, NKBIA, NAMPT and enrichment of REACTOME pathways associated with neutrophil degranulation as well as proteins CD35, CD55, CLEC12A and CD48 (Figure 2G, Supplementary Figure 2C). This reflects a broader limitation of sc-RNAseq, which is suboptimal for profiling neutrophils due to their low RNA content and fragility. In contrast, methods such as flow cytometry, which better preserve granulocyte populations in fresh samples, offer a more reliable approach for neutrophil analysis, as demonstrated in our recent study of CF BAL ^31^. Overall, our CF and control paediatric BAL atlas resulted in >190,000 cells from 45 samples being classified into 43 cellular subtypes.

### 3. Widespread dysregulation of BAL macrophage responses in preschool children with CF compared to non-CF controls

We firstly compared BAL immune cell proportions between children with CF (not on any modulator therapy) (CF (no mod)) (n=25) and non-CF controls (n=8). Given that out BAL samples are comprised of >80% macrophages, we examined macrophages subtype proportions as a fraction of total macrophages, whilst proportions of T/NK and rare cells were examined relative to the total pool of T/NK and rare cells. This revealed that children with CF have a significantly reduced proportion of monocytes (CF (no mod)=0.0102, non-CF control=0.0248; p=0.009, FDR=0.001), increased proportion of NK-T cells (CF (no mod)=0.0042, non-CF control=0.0025; p=0.01, FDR=n.s.), increased CD4 Tregs (CF (no mod)=0.0173, non-CF control=0.0102; p=0.01, FDR=n.s.), and increased macro-IGF1 cells (CF (no mod)=0.0506, NON_CF.CTRL=0.0234; p=0.001, FDR=0.0233) relative to non-CF controls (Figure 3A).

**Figure 3.**
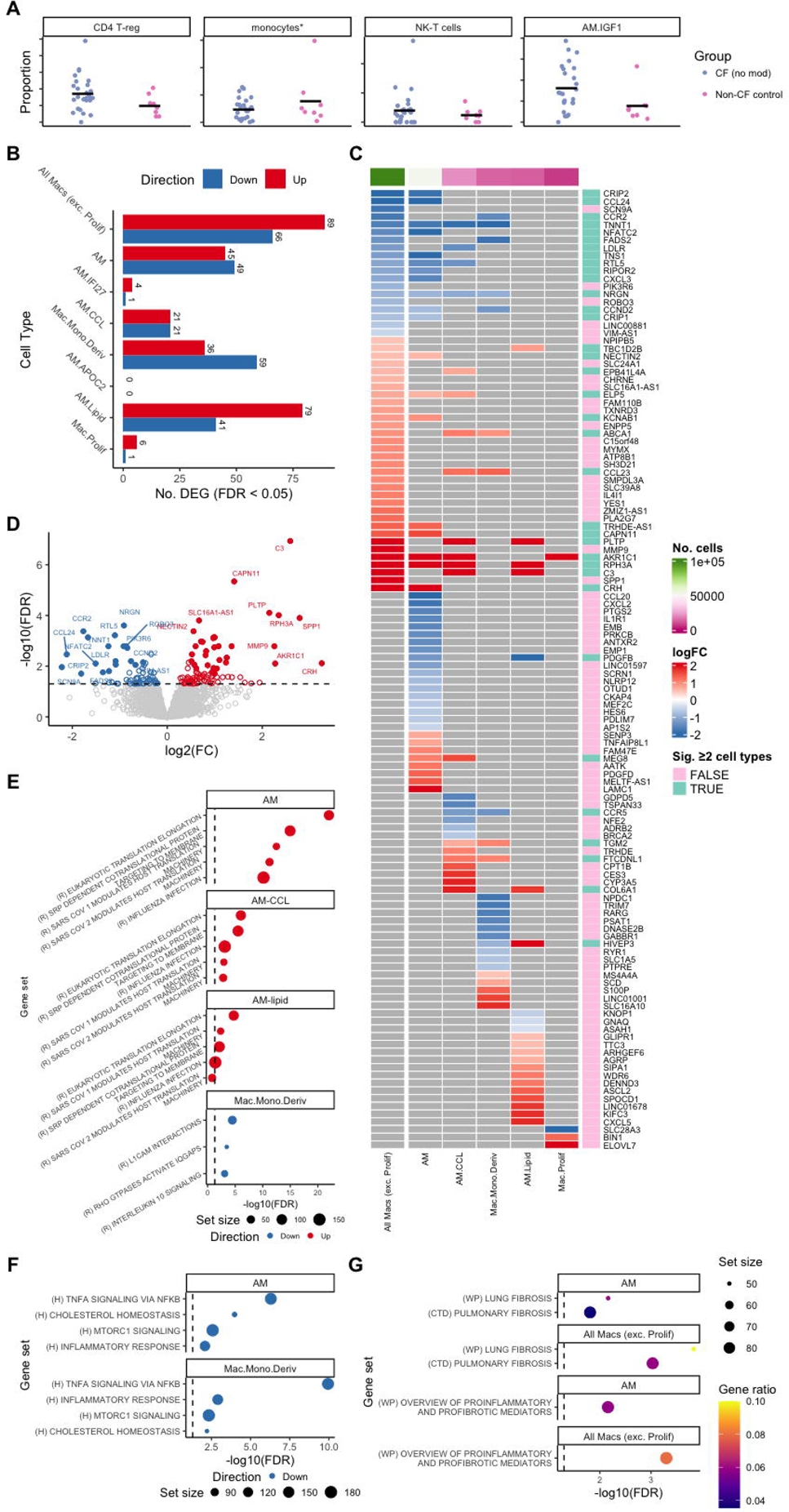
Widespread dysregulation of BAL macrophage responses in preschool children with CF compared to non-CF controls. (A) Proportions of CD4 Tregs, monocytes, NK-T cells, and macro-IGF1 cells in BAL samples from children with CF (no modulator) (n=25) and non-CF controls (n=8). *indicates significant FDR adjusted p-value. (B) Summary of the number of differentially expressed genes between CF (no modulator) (CF (no mod)) and non-CF control groups for each tested macrophage subpopulation. (C) Heatmap depicting the most significant differentially expressed genes between CF (no modulator) (CF (no mod)) and non-CF control groups for each tested macrophage subpopulation. (D) Volcano plot depicting up- and down-regulated genes in the CF (no modulator) (CF (no mod)) and non-CF comparison within the total macrophage pool. (E) REACTOME pathway analysis of differentially expressed genes within the macro-alveolar, macro-CCL, macro-lipid and macro-monocyte-derived populations. (F) HALLMARK pathway analysis of differentially expressed genes within the macro-alveolar and macro-monocyte-derived populations. (G) Lung fibrosis pathway analysis of differentially expressed genes within the total macrophage and macro-alveolar populations. WP = Wiki Pathways; CTD = Comparative Toxicogenomics Database.

To identify gene expression programs associated with disease we performed differential expression analysis within cell types across patient groups using a pseudo-bulk approach (see methods). All DEGs across all subpopulations are provided on the analysis website (https://github.com/Oshlack/paediatric-cf-inflammation-citeseq/tree/main/output/dge_analysis). We identified 518 differentially expressed genes (DEGs) between samples from children with CF (no mod) and non-CF controls within alveolar macrophages, monocyte-derived macrophages, macro-CCL, and macro lipid (Figure 3B). A heatmap depicting the most significant DEGs for each macrophage subpopulation is shown in Figure 3C, and a representative volcano plot depicting up and down regulated genes within the total macrophage pool is shown in Figure 3D. Enrichment analysis of these DEGs using the REACTOME and HALLMARK databases revealed upregulation of pathways associated with “eukaryotic translation and elongation” and “SRP dependent co-translational protein targeting to membrane”, in macro-alveolar, macro-CCL and macro-lipid populations from children with CF, which may be related to CFTR protein misfolding seen in Phe508del CFTR variants ^32–34^ (Figure 3E). Conversely, downregulation of pathways associated with TNFA signalling, inflammatory responses, MTORC1 signalling, and cholesterol homeostasis were observed in both alveolar and monocyte-derived macrophages from children with CF (Figure 3F). Monocyte-derived macrophages from children with CF showed downregulation of pathways not observed in other macrophage subpopulations including IL-10 signalling. Interestingly, IL-10 protein levels have been shown to be reduced in people with CF ^17^.

We also observed transcriptional signatures that suggest remodelling of pulmonary fibrosis pathways in CF alveolar macrophages. This was evident in multiple gene sets, including “lung fibrosis” (FDR=0.0001), “pulmonary fibrosis ctd” (FDR=0.0009), and “overview of proinflammatory and profibrotic mediators” (FDR=0.0005) (Figure 3G).

Due to their abundance in BAL, macrophage subpopulations provided the most power to detect differential gene expression and were therefore the focus of our analysis. However, we also analysed differential expression within other cell types in BAL where cell numbers permitted (CD4 T cells, CD8 T cells, and dendritic cells). This showed limited differences between children with CF and controls, with five genes differentially expressed in both CD8 T cells and dendritic cells, the majority of which associated with pro-inflammatory effects and upregulated in the CF group, and none differentially expressed in CD4 T cells (Supplementary Figure 3).

### 4. BAL macrophage transcriptional signatures are associated with CF bronchiectasis

Within the CF (no mod) samples, 8 had evidence of bronchiectasis on chest CT and 17 had no evidence of bronchiectasis on chest CT. We therefore compared BAL cell proportions and differential gene expression between these two groups, where those with bronchiectasis were termed “severe” and those without bronchiectasis were termed “mild”. Samples from children with mild lung disease had elevated proportions of CD4 T-IFN cells (mild=0.01439, severe=0.0057; p=0.01, FDR=n.s.) and reduced proportions of HSP+ B cells (mild=0.0026, severe=0.0091; p=0.03, FDR=n.s.) relative to samples from children with severe lung disease (Figure 4A). As these cell populations have not been observed in people with CF before, the biological significance of this finding is unclear.

**Figure 4.**
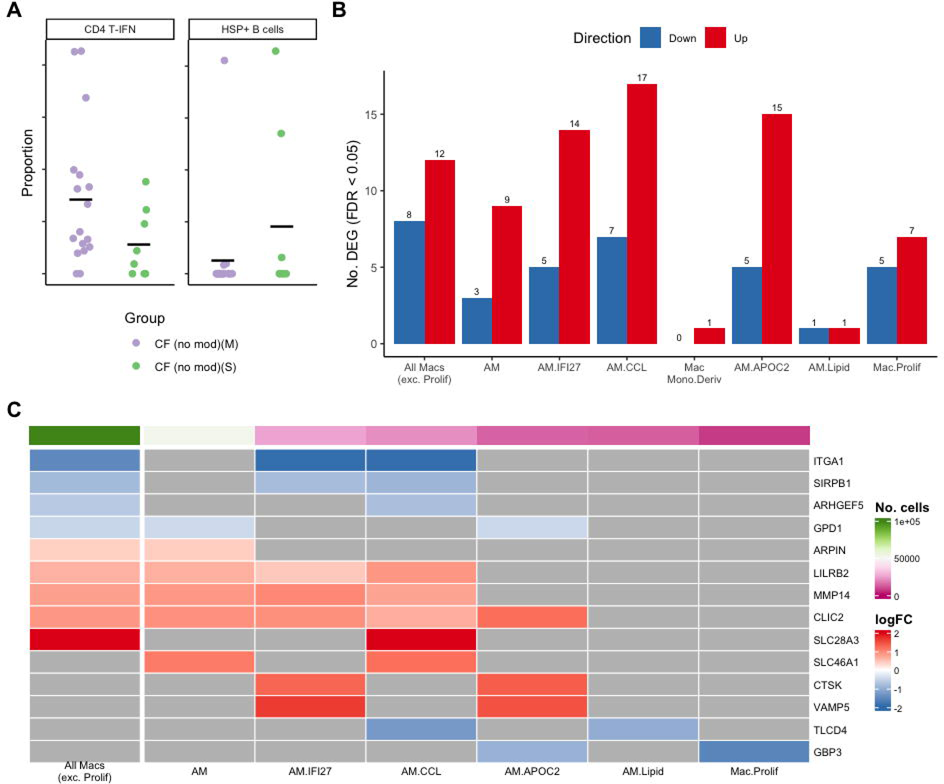
BAL macrophage transcriptional signatures are associated with CF bronchiectasis. (A) Proportions of CD4 T-IFN and HSP+ B cells in BAL samples from CF children with bronchiectasis (CF (no mod)(S) = “severe”) (n=8) and no bronchiectasis (CF (no mod)(M) = “mild”) (n=17). (B) Summary of the number of differentially expressed genes between CF severe and CF mild groups for each tested macrophage subpopulation. (C) Heatmap depicting genes that were differentially expressed in multiple macrophage subpopulations between CF severe and CF mild groups.

When exploring differential gene expression within macrophage subpopulations, we show key differences between mild and severe lung disease within the alveolar, macro-CCL, macro-IFI27, macro-APOC2, and proliferating macrophage populations (Figure 4B, Supplementary Figure 4). In total, we identified 110 DEGs, several of which overlapped across multiple macrophage subpopulations, suggesting these genes are playing key roles in the response by these cells (Figure 4C). These included *CLIC2, SLC28A3, MMP14, VAMP5, CTSK, LILRB2, SLC46A1* and *ARPIN* which were upregulated across multiple macrophage subpopulations in severe lung disease compared to mild lung disease, and *ARHGEF5, GPD1, TLCD4, GBP3, SIRPB1* and *ITGA1* which were downregulated in multiple macrophage subpopulations in severe lung disease compared to mild lung disease (Figure 4C). Some of the upregulated genes (CLIC2, MMP14, CTSK) have been associated with pulmonary fibrosis ^35^ and LILRB2 is an inhibitor of macrophage activation and upregulation of this gene further highlights macrophage dysfunction in CF ^36^.

As in section 3, differential expression analysis within CD4 T cells, CD8 T cells and dendritic cells in children with CF with bronchiectasis (severe) and those without (mild) was performed and summarised in Supplementary Figure 5. This showed that *AREG* expression was reduced in CD4 T cells from children with bronchiectasis. The relationship between AREG and lung damage is complex, in some situations it promotes tissue repair after lung injury and in others lung diseases it is associated with fibrosis ^37^. *IER2*, *MSMO1*, and *FBLN5* were reduced in CD8 T cells from children with bronchiectasis. Conversely, *CMC1* (a marker of CD8 T cell activation)^38^ and *CST7* expression was upregulated in CD8 T cells from children with bronchiectasis. Finally, *FABP4* and *SCD* expression (both associated with lipid metabolism) was upregulated in dendritic cells from children with bronchiectasis.

### 5. The CFTR modulator ivacaftor modifies BAL macrophage response in early life

We also explored the effect of CFTR modulator therapy on early life lung inflammation by comparing responses in samples from CF children on two modulator therapies: ivacaftor (8 BAL samples) and lumacaftor/ivacaftor (4 BAL samples). Relative to CF(no mod) (n=25), samples from children on ivacaftor (CF (iva)) have reduced proportions of NK-T cells (CF (iva)=0.0012, CF(no mod)=0.0042; p=0.01, FDR=n.s.) (Figure 5A), which were elevated in CF (no mod) relative to non-CF controls (Figure 3A). CD4 T-IFN cells, associated with mild lung disease in our cohort (Figure 4A), tended to be elevated in CF (iva) relative to non-CF controls (CF (iva)=0.0222, CF (no mod)=0.0116; p=0.05, FDR=n.s.) (Figure 5A). Monocytes, which were reduced in CF (no mod) relative to non-CF controls (Figure 3A), remained reduced following treatment with ivacaftor (CF (iva)=0.1032, CF (no mod)=0.0990; p=0.02, FDR=n.s.) (Figure 5A). When exploring proportion differences induced by treatment with lumafactor/ivacaftor (CF (luma/iva) we did not see a reduction in NK-T cells as we did for CF (iva) (CF (no mod)=0.0232, CF (luma/iva)=0.0014; p=0.89, FDR=n.s.) (Figure 5B). CD4 Tregs, which we showed earlier to be elevated in CF (no mod) relative to non-CF controls (Figure 3A), remained elevated in CF (luma/iva) (CF (luma/iva)=0.0394, CF (no mod)=0.0173; p=0.03, FDR=n.s.), and monocytes tended to remain reduced (CF (luma/iva)=0.1150, CF (no mod)=0.0990; p=0.09) (Figure 5B). Similar to CF (iva), CD4-IFN cells tended to be elevated in CF (luma/iva) relative to non-CF controls (CF (luma/iva)=0.0095, CF (no mod)=0.0116; p=0.06, FDR=n.s.). Relative to CF (no mod), as well as compared to non-CF controls, we showed elevated proportions of the macro-CCL18 subset in CF (luma/iva) which was not observed in CF (iva) (CF (luma/iva)=0.1045, CF (no mod)=0.0751, non-CF control=0.0608; p=0.01 and p=0.02, respectively, FDR=n.s.) (Figure 5B).

**Figure 5.**
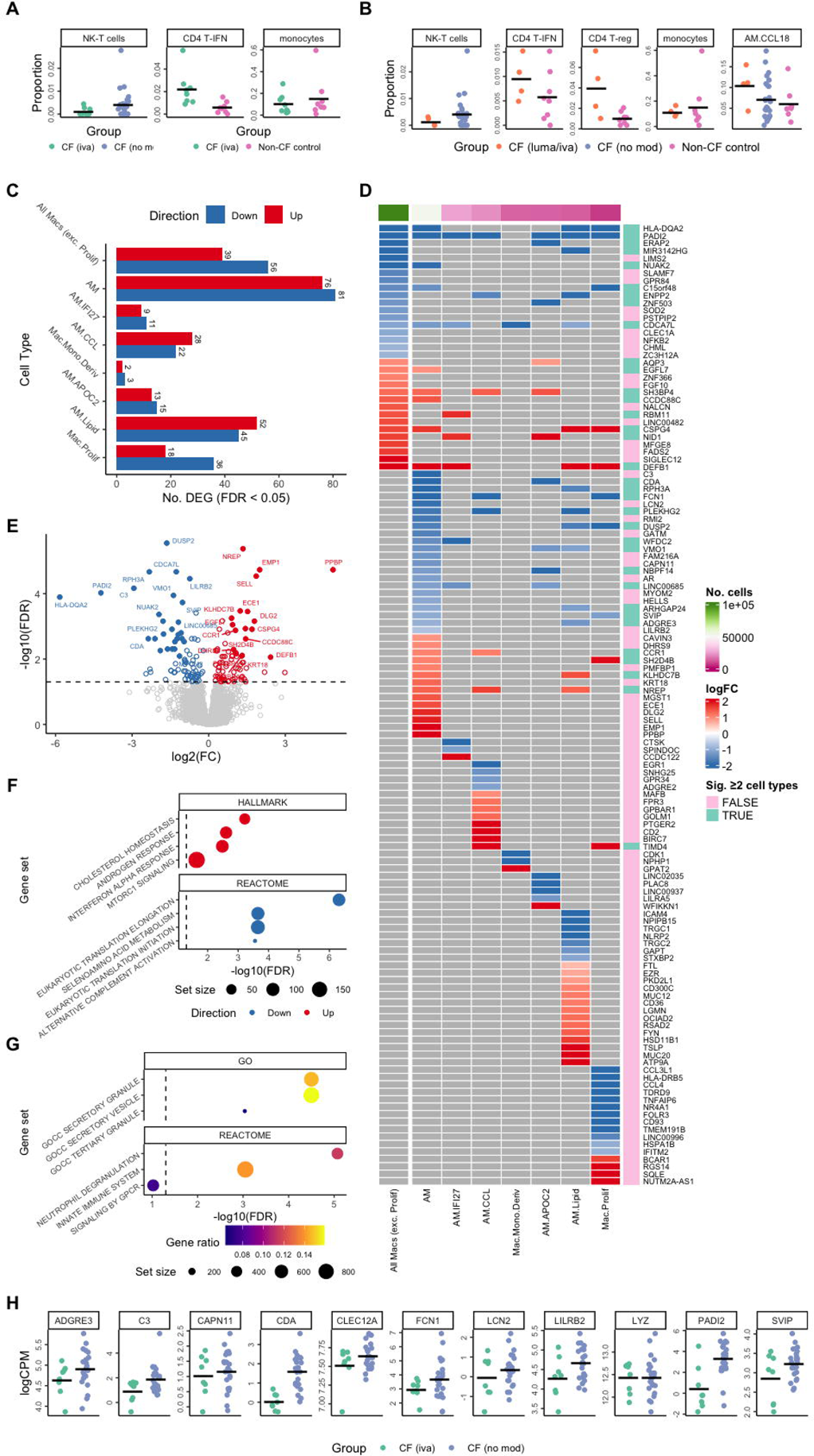
The CFTR modulator ivacaftor modifies BAL macrophage function in early life. (A) Proportions of NK-T cells, CD4 T-IFN, and monocytes in BAL samples from children treated with ivacaftor (CF (iva)) (n=8), CF (no mod) (n=25) and non-CF controls (n=8). (B) Proportions of NK-T cells, CD4 T-IFN, CD4 T-reg, monocytes and macro-CCL in BAL samples from children treated with lumacaftor/ivacaftor (CF (luma/iva)) (n=4), CF (no mod) (n=25) and non-CF controls (n=8). (C) Summary of the number of differentially expressed genes between CF (iva) (n=8) and CF (no modulator) (CF (no mod)) (n=25) for each tested macrophage subpopulation. (D) Heatmap depicting the most significant differentially expressed genes between CF (iva) and CF (no modulator) (CF (no mod)) groups for each tested macrophage subpopulation. (E) Volcano plot depicting up- and down-regulated genes in the CF (iva) and CF (no modulator) (CF (no mod)) comparison within the total macrophage pool. (F) HALLMARK and REACTOME pathway analysis of differentially expressed genes within the macro-alveolar population. (G) Gene ontology and REACTOME pathway analysis revealing remodelling of neutrophil response pathways in the macro-alveolar population. (H) Selected differentially expressed genes related to neutrophil pathways within the macro-alveolar population in CF (iva) and CF (no modulator) (CF (no mod)) comparison (all FDR p<0.05).

Differential expression analysis within macrophage populations revealed a significant effect of ivacaftor treatment (CF (iva)) on macrophage gene expression relative to CF (no mod) (506 DEGs in total) (Figure 5C-E). Conversely, we observed no significant differences in gene expression between lumacaftor/ivacaftor (CF (luma/iva)) samples and CF (no mod) samples in any macrophage subpopulation, suggesting a limited effect of lumacaftor/ivacaftor on macrophage function in the CF airway. We note however the more limited numbers in the CF (luma/iva) group (n=4) relative to the CF (iva) group (n=8).

Enrichment analysis of CF (iva) vs CF (no mod) DEGs using the REACTOME and HALLMARK gene sets revealed upregulation of pathways associated with cholesterol homeostasis, androgen responses, and IFN-A responses in CF (iva) relative to CF (no mod)across these macrophage subpopulations. Many of these pathways were shown to be impaired and downregulated in CF children not on modulator therapy compared to non-CF controls (Figure 3) suggesting that the treatment restores some of the lost function. Further, eukaryotic translation was downregulated in CF (iva) relative to CF (no mod) across these macrophage subpopulations. These pathways were upregulated in CF children not on modulator therapy compared to non-CF controls (Figure 3). Figure 5F shows representative pathway analysis within the alveolar macrophage population, however similar responses were also observed in other macrophage subpopulations including macro-CCL and macro-lipid (see analysis website). These findings suggest that ivacaftor treatment restores some of the CF macrophage-associated functional deficits in early life.

Ivacaftor also induced remodelling of neutrophil response pathways in alveolar macrophages as highlighted by enrichment of the Gene Ontology pathways secretory granules (FDR=0.00003, 23 DEGs) and secretory vesicles (FDR=0.00003, 25 DEGs) and the REACTOME pathway neutrophil degranulation (FDR=8.45E-06, 18 DEGs) (Figure 5G). This signature was unique to the alveolar macrophage subpopulation, highlighting a potential key role for these cells in regulating the neutrophil response following initiation of ivacaftor. Key genes in these pathways are shown in Figure 5H, all of which were significantly downregulated (all FDR<0.05) in CF (iva) samples relative to CF (no mod) samples.

### 6. Residual abnormalities between CF ivacaftor and controls

As a further avenue to determine the effects of ivacaftor treatment on CF macrophage associated functional deficits, we compared cell-specific gene expression in CF (iva) with that of non-CF controls and contrasted that to the CF (no mod) vs. non-CF control comparison. Twelve genes were significantly differentially expressed in both comparisons, with highly correlated log fold changes, suggesting similar dysregulation in both ivacaftor treated and untreated children with CF relative to the non-CF controls. Of these 12 genes, *IL6, CXCL3, IL1B and TNS1* are downregulated in the CF samples for both comparisons, whilst Y*ES1, TRHDE, ARL14EPL, MELTF, PPIF, ITSN1 and EPB41L4A* were all upregulated (Figure 6A). Of note, *PPIF* has been shown to be upregulated in CF respiratory epithelium when compared to controls ^39^, and *YES1* interacts with CFTR proteins that have been rescued by modulator treatment and promotes shorter half-life of rescued CFTR ^40^. Fifty-seven genes were uniquely differentially expressed between CF (iva) and non-CF controls (Figure 6A), including increased expression of *ADAMTS1*, which has been associated with reduced airway neutrophilia, reduced airway remodelling and improved lung function in other airway diseases^41^. The Hallmark gene sets IL2 STAT5 signalling, TNFA via NFKB signalling, and inflammatory response are amongst the most significantly over-represented within the DEGs when comparing CF (no mod) versus non-CF controls, but only TNFA via NFKB signalling and inflammatory response remain significant for CF (iva) vs. non-CF controls (Figure 6B). The IL2 STAT5 pathway is associated with the development of Th2 inflammation ^42,43^, which is implicated in CF related complications such as allergic bronchopulmonary aspergillosis. This finding suggests ivacaftor treatment may reduce the development of Th2 inflammation via IL2 STAT5 signalling. Similar to these results, in a recent analysis of BAL cytokine concentration, we demonstrated elevated Th2 inflammation in children with CF not on a modulator, but no evidence of Th2 inflammation in children treated with ivacaftor ^31^. The observation that gene sets for TNFA via NFKB signalling and inflammatory response remain significant for CF (iva) vs non-CF controls suggests that while ivacaftor is associated with improvements in some pathways, the inflammatory environment in the lung remains different to controls.

**Figure 6.**
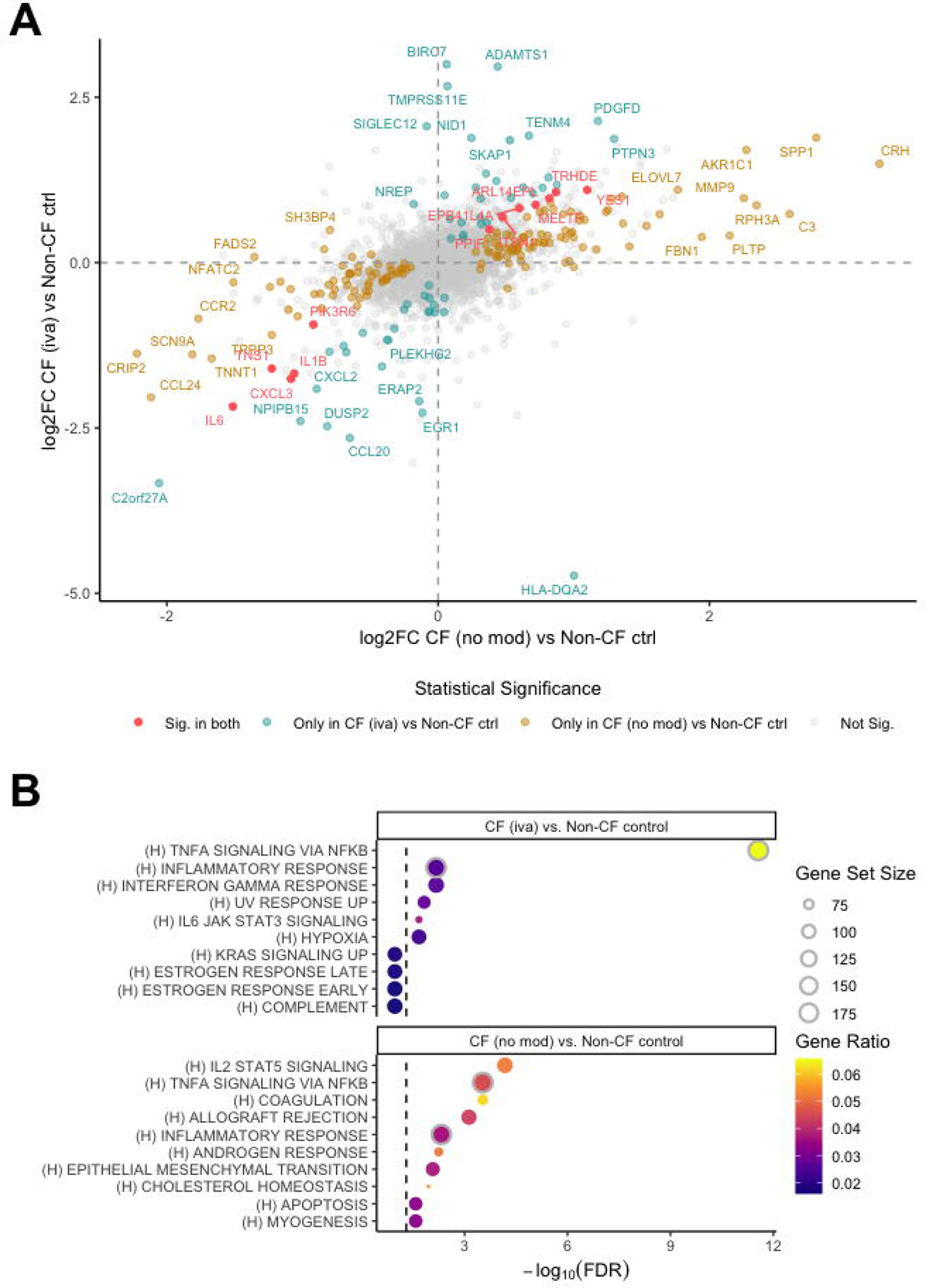
Residual inflammation in CF ivacaftor relative to controls. Comparison of differential gene expression results for samples from children treated with ivacaftor (CF (iva)) (n=8) versus non-CF control (n=8) and CF (no modulator) (CF (no mod)) (n=25) versus non-CF control (n=8) samples in the overall macrophage population. (A) Difference in gene expression between CF (no mod) versus non-CF control compared to CF (iva) versus non-CF control. Genes that were significantly differentially expressed in both comparisons are highlighted in pink; genes that were only significant in CF (iva) versus non-CF control are highlighted in teal; genes that were only significant in CF (no mod) versus non-CF control are highlighted in mustard; genes that were not significant in either comparison are grey. (B) The top 10 Hallmark gene sets that were found to be overrepresented amongst significantly differentially expressed genes when comparing CF (iva) versus non-CF control (top panel) and CF (no mod) versus non-CF control (bottom panel). Significant Hallmark gene sets that are in the top 10 for both comparisons are highlighted by a grey circle outline. The gene set size is the total number of genes in the gene set; the Gene Ratio is the numbers of DEGs in the gene set divided by the gene set size.

### 7. CF-associated genes show differential expression by disease status and ivacaftor treatment in epithelial cell subsets

As our dataset contained only 1,847 epithelial cells, including very few CFTR-expressing cells (Supplementary Figure 6A), our ability to detect novel DEGs or make definitive statements about CFTR expression is limited (Supplementary Figure 6B). Thus, we leveraged findings from prior large-scale studies (>40,000 epithelial cells from 19 CF patients and 19 controls). Sun and Zhou (2025)^44^ and Carraro et al. (2021)^4^ both reported DEGs between CF and control airway epithelial cells using different analytical strategies. Testing the combined set of their DEGs in our data, we found significant differential expression (FDR < 0.05) for 20 genes in ciliated epithelial cells (Supplementary Figure 7A) and 26 in secretory cells (Supplementary Figure 8A) between non-CF controls and CF (no mod). Furthermore, six of their DEGs were also significantly different between CF (iva) and CF (no mod) in ciliated cells (Supplementary Figure 7B), and four in secretory cells (Supplementary Figure 8B). This included increased expression of *SAA1* and *SAA2* in ciliated cells in the CF (no mod) samples relative to both non-CF controls and CF (iva). *SAA1* and *SAA2* are associated serum amyloid A, a general biomarker of inflammation, which is increased in people with CF. ^45^ *KRT19*, which helps to stabilise CFTR at the cell surface and is reduced in *in vitro* models of CF ^46^, was increased in secretory cells from non-CF controls, and the CF (iva) group relative to CF (no mod).

## DISCUSSION

In this study, we integrated transcriptome-wide single-cell RNA sequencing data, highly multiplexed surface protein expression, and functional pathway analysis to extensively characterise 43 immune and epithelial cell populations in early life BAL. We then analysed this extensive BAL atlas to compare transcriptional responses in samples from children with CF and non-CF controls and discovered widespread dysregulation of macrophage function in the early life CF lung, which was further altered by the early development of bronchiectasis. We showed that the CFTR modulator ivacaftor restores some of these macrophages related functional deficits, and that the modulator lumacaftor/ivacaftor was not associated with any detectable change in transcriptional response.

To robustly detect differences in gene expression between sample groups within a cell type, scRNA-seq analysis must account for both cell-to-cell and sample-to-sample variability ^19,47^. Failure to model inter-individual variation can lead to inflated false discovery rates, as shown by Squair et al. (2021) ^19^, who demonstrated that many commonly used single-cell differential expression methods, which were primarily designed for identifying cluster-specific marker genes, do not adequately control for sample-level effects. Our analysis adopts best-practice pseudobulk differential expression analysis, using RUVSeq ^48^ in combination with edgeR ^49^ to remove unwanted sources of variation. This represents a novel application of RUVSeq in the single-cell context, enabling more accurate modelling of biological and technical variability, and improving the reliability of our gene-level inferences.

Previous scRNA-seq studies involving people with CF have largely characterised adult samples and primarily focused on respiratory epithelium rather than immune cells. This is also true for the broader pulmonary scRNA-seq field ^8,21^. Our paediatric-specific dataset revealed several novel cell subtypes not observed in adult studies, such as CD4 T cells with NFκB and IFN activation signatures. As these cells were found in both children with CF and non-CF controls, they are unlikely to be disease specific and may represent cell types specific to early life, a finding which needs external validation. The proportion of broad cell types identified differs from previous studies in that there is a low proportion of neutrophils in our work, likely due to the impact of cryopreservation ^50^, along with the known difficulties in detecting neutrophils using droplet based sc-RNAseq technology.

We showed dysregulation of a range of immune response pathways in four key macrophage populations (macro-alveolar, macro-monocyte derived, macro-CCL and macro-lipid) in children with CF relative to age-matched controls. This included upregulation of pathways associated with immune defence, downregulation of pathways associated with TNF/IFN signalling and cholesterol homeostasis, as well as evidence for remodelling of pulmonary fibrosis pathways. A recent study applying scRNAseq to lungs from CF and wildtype piglets revealed similar alterations in CF lung myeloid cell gene expression, including elevated pathways associated with immune defence response, reduced cytoplasmatic translation, and enrichment for S100 alarmins and matrix metalloproteinases (MMPs) ^51^. This was also associated with impaired phagocytosis function, similar to previous work in sputum from adults with CF ^5^.

In our study, macrophages from children with evidence of bronchiectasis had higher expression of genes such as *CLIC2, MMP14* and *CTSK*, which have been associated with pulmonary fibrosis and indeed the MMP proteins have been explored as therapeutic targets in CF lung disease ^35,52,53^. We also showed upregulation of *LILRB2,* a negative regulator of myeloid cell activation, in early life CF bronchiectasis. Recent work has shown anti-LILRB2 therapy increases macrophage activation (*NFKB* and *STAT1*) as well as alters genes involved in lipid and cholesterol metabolism ^36^. Overall, our findings and others highlight that in those not eligible for modulator therapy, where gene therapy is the next therapeutic approach being trialled, CFTR may need to be corrected in macrophages as well as the respiratory epithelium to prevent the harms of CF-related pulmonary inflammation.

Alveolar macrophages from the ivacaftor treated group had reduced expression of genes associated with neutrophil recruitment and degranulation relative to the CF-no modulator group. Supporting this, we have previously shown ivacaftor is associated with a reduced proportion of neutrophils and reduced concentration of neutrophil-related cytokines in BAL ^31,54^, and elexacaftor/tezacaftor/ivacaftor (ETI) therapy was also recently associated with reduced expression of neutrophil recruitment pathways in macrophages obtained from nasal brushings ^7^. Together these findings suggest that highly effective modulator therapy, commenced in early life, may be effective in preventing the development of neutrophil dominant inflammation. We also showed in several macrophage subsets that pathways observed to be downregulated in CF-no modulator compared with non-CF controls were upregulated by ivacaftor, including cholesterol homeostasis and interferon-A responses, indicative of an ivacaftor-induced restoration of key macrophage functions similar to that observed with ETI ^7^. We showed no detectable effect of lumacaftor/ivacaftor treatment when compared to children not on modulator therapy, although we acknowledge the smaller number of participants in this cohort. This limited impact of lumacaftor/ivacaftor therapy is perhaps not surprising given the modest clinical impact of this treatment when compared to other modulator therapies ^55^.

While we showed evidence of improvements in macrophage function in the ivacaftor treated group relative to the CF-no modulator group, we also observed residual differences between the CF (iva) group and non-CF controls. This included altered airway inflammation with downregulation of gene pathways related to TNFA signalling and inflammatory response, observed in both CF (no mod) and CF (iva) groups. The downregulation of TNFA signalling in BAL macrophages was at first surprising, given that TNF and other inflammatory protein levels are higher in BAL fluid of children with CF ^31^. This suggests that other cell types not captured in our data, such as infiltrating neutrophils, are the likely source of TNF and related inflammatory proteins in the CF airways. Supporting this, we have recently shown a strong positive correlation between these protein levels (including TNFα, IL-6, IL-1β, and IL-8) and neutrophil cell count in CF BAL^31^. Previous work has shown that monocyte-derived macrophages, sampled from the nose and blood, from older children and adults with CF show elevated inflammatory signalling relative to controls, with conflicting evidence as to the effect of CFTR modulators on this response ^7,56^. The downregulation of these pathways in macrophages in our data may be unique to early life or to the lower airway, and needs validation in other studies.

This study has limitations which must be acknowledged. The cross-sectional design limits the ability to infer causality with regards to treatment effect. Further, participants treated with modulators were treated for different lengths of time, however as modulator therapy is associated with changes in CFTR function and clinical outcomes within 2 weeks of treatment initiation this is unlikely to be significant ^57^. New modulators, such as ETI and now vanzacaftor–tezacaftor–deutivacaftor, mean that ivacaftor and lumacaftor/ivacaftor have been superseded. However, at the time of this study these were the agents used in clinical practice for children in Australia and the results still offer insights into the degree of CFTR function correction needed to alter pulmonary inflammation. The fact that the control group is made up of children undergoing surgery for upper airway pathology is another limitation that reflects the difficult in obtaining BAL samples from children for research. To try and ensure the control group had no lung disease we used a parent questionnaire and excluded any participants who reported lung pathology.

In summary, this is the most comprehensive, multi-omic single-cell analysis of bronchoalveolar lavage from preschool children. These data have identified novel cell populations in the early life lung, highlighted multiple aberrant pathways leading to macrophage dysfunction in preschool CF, and demonstrated the benefits of the CFTR modulator ivacaftor at single-cell level.

## METHODS

### Study participants

This study included 45 bronchoalveolar (BAL) samples from children with CF (n=37) and non-CF controls (n=8) (Extended Data File 1). The CF group consisted of children who had surveillance bronchoscopy as part of clinical care through the AREST-CF program ^58^), and the control group were children without cystic fibrosis who underwent general anaesthesia for assessment of upper airway disease (such as stridor). Children had bronchoscopy performed at the time of clinical stability with no clinical features of acute infection. The median age of the CF participants was 3.03 years (range 0.5-6.2 years) and the median age of the non-CF controls was 2.11 years (range 0.84-8.4 years). Within the CF samples, n=25 were not on any CFTR modulator therapy, 8 were receiving the CFTR modulator ivacaftor and 4 were receiving the CFTR modulator lumacaftor/ivacaftor.

### Ethics statement

This study was reviewed and approved by Royal Children’s Hospital Research Ethics Committee (HREC #25054). Written informed consent to participate in the studies was provided by the participants’ legal guardian/next of kin.

### Bronchoalveolar lavage procedure and processing of samples

Three aliquots of sterile normal saline (1mL/kg, max 20mL per aliquot) were instilled into the right middle lobe and another lavage of an aliquot was performed in the lingula or the site of greatest disease. This study used BAL from aliquots two and three from the right middle lobe. BAL was assessed for infection as per standard clinical testing in the RCH clinical microbiology laboratory. BAL samples were processed according to our openly available protocols ^59^. In brief, samples were immediately placed on ice and processed within 1 hour of the procedure. Samples were centrifuged at 300 x g for 7 min at 4°C. The cell pellet was resuspended in 10mL of media (RPMI supplemented with 2% fetal calf serum (FCS)), filtered through a 70uM filter and centrifuged at 300 x g for 7 min at 4°C. Supernatant was discarded and the cell pellet resuspended in media for cell counting. The sample was then centrifuged at 300 x g for 7 min and resuspended in equal parts media and chilled freezing solution (FCS with15% dimethyl sulfoxide (DMSO)). The freezing solution was added to resuspended cells drop by drop on ice. The samples were transferred to cryovials, and then cooled at −1°C per minute to −80 °C overnight before being transferred to liquid nitrogen for storage.

### Collection of clinical information

Medication records were reviewed, and participants were classified as being treated or not treated with CFTR modulator therapy. At the time of the study, modulator therapies that were available to the participants were ivacaftor (iva) (for children with an eligible class III or IV mutation), and lumacaftor/ivacaftor (luma/iva) (for children homozygous for Phe508 del mutation). Children not on modulator therapy either had a genotype ineligible for modulator therapy, or their parents had elected not to start treatment. To assess lung disease severity, each participants most recent computed tomography scan was reviewed by a clinician trained in the assessment of early life CF scans, and participants were classified as having bronchiectasis (herein denoted severe lung disease) or not (herein denoted mild lung disease). As per previous studies, bronchiectasis was defined as a bronchus-to-artery ratio of more than 1.0 or the presence of a non-tapering bronchus in the transverse plane ^58^. This classification was made after a review of the participants most recent computed tomography scan, by a clinician trained in the review of paediatric CF scans who was blinded to the participants clinical status.

### Processing of BAL samples for single-cell sequencing

Cryopreserved BAL cells were thawed in 10mL media (RPMI supplemented with 10% heat-inactivated FCS) with 25U/mL benzonase at 37°C and centrifuged at 300 x g for 10 min. The pellet was resuspended in 1mL PBS for cell counting. Following cell count, 9mL PBS was added to the tube and cells were centrifuged at 300 x g for 10 min. Supernatant was discarded and the cell pellet resuspended in PBS for viability staining using live/dead fixable near infrared viability dye (Invitrogen) according to manufacturers’ instructions. The viability dye reaction was stopped by the addition of FACS buffer (2% heat-inactivated FCS in PBS 2mM EDTA) and cells were centrifuged at 400 x g for 5 min. Cells were resuspended in FACS buffer for live, single cell sorting using a BD FACSAria Fusion according to the gating strategy outlined in Supplementary Figure E1. Samples were processed for single-cell sequencing in 6 batches, the first batch (samples 40.1-43.1) were processed for single-cell RNA sequencing (without the addition of ADT’s) using the “scRNAseq protocol” below, and the remaining 5 batches (samples 1.1-39.2) were processed for CITE-seq using the “CITEseq protocol” below.

### scRNA-seq protocol

Sorted cells were counted and assessed for viability with Trypan Blue using a Countess automated counter (Invitrogen) and then resuspended at a concentration of 800-1000 cells/μL (8×105 to 1×106 cells/mL). Final cell viability estimates ranged between 92-96%. Single cell suspensions were loaded onto 10X Genomics Single Cell 3’ Chips along with the reverse transcription (RT) mastermix as per the manufacturer’s protocol for the Chromium Single Cell 3’ Library (10X Genomics; PN-120233), to generate single cell gel beads in emulsion (GEMs). Reverse transcription was performed using a C1000 Touch Thermal Cycler with a Deep Well Reaction Module (Bio-Rad) as follows: 55°C for 2h; 85°C for 5min; hold 4°C. cDNA was recovered and purified with DynaBeads MyOne Silane Beads (Thermo Fisher Scientific; Cat# 37002D) and SPRIselect beads (Beckman Coulter; Cat# B23318). Purified cDNA was amplified as follows: 98°C for 3min; 12x (98°C for 15s, 67°C for 20s, 72°C for 60s); 72°C for 60s; hold 4°C. Amplified cDNA was purified using SPRIselect beads and sheared to approximately 200bp with a Covaris S2 instrument (Covaris) using the manufacturer’s recommended parameters. Sequencing libraries were generated with unique sample indices (SI) for each chromium reaction. Libraries for all samples were multiplexed and sequenced across on 2×150 cycle flow cells on an Illumina NovaSeq 6000 (26bp (Read 1), 8bp (Index), and 98 bp (Read 2)).

The Cell Ranger Single Cell Software Suite (version 6.0.2) by 10x Genomics was used to process raw sequence data into FASTQ files and read counts per cell, per gene. First, raw base calls from multiple flow cells were demultiplexed into separate pools of samples. Reads from each pool were then mapped to the GRCh38/hg38 genome (version 12) using STAR.

### CITE-seq protocol

Sorted live single cells were centrifuged at 400 x g for 5min at 4°C and resuspended in 25μL of cell staining buffer (BioLegend). Human TruStain FcX FC blocking reagent (BioLegend) was added according to manufacturers’ instructions for 10min on ice. Each tube was made up to 100μL with cell staining buffer and TotalSeq Hashtag reagents (BioLegend) were added to each sample for 20min on ice. Cells were washed with 3mL cell staining buffer and centrifuged at 400xg for 5min at 4°C. Supernatant was discarded and each sample resuspended at 62,500 cells/100μL following which 100μL of each sample were pooled into one tube. Pooled cells were centrifuged at 400xg for 5min at 4°C, supernatant discarded, and resuspended in 25μL cell staining buffer and 25ul of TotalSeqA Human Universal Cocktail v1.0 (BioLegend) for 30min on ice. Cells were washed in 3mL cell staining buffer and centrifuged at 400xg for 5min at 4°C. Following two more washes, cells were resuspended in PBS + 0.04% BSA for Chromium captures. Single-cell captures and library preparations were processed with the 10x Genomics Chromium single-cell Platform using the 10x Chromium Next GEM single-cell 3’ Reagent V3.1 Dual Index kits (10x Genomics, USA) following the manufacturer’s manual, where each pool was run on two captures to maximise cell numbers. The barcoded cDNA was PCR-amplified and scRNA-seq libraries were constructed using the 10x 3’v3.1 library kits. The TotalSeq-A scADT-seq and scHTO-seq libraries were constructed as per manufacturer’s instructions (BioLegend). The duplicate scRNA-seq, scADT-seq and scHTO-seq libraries were quantified by Tapestation 4200 D1000 chip (Agilent). Upon input normalisation, the libraries were pooled and sequenced on the Illumina NextSeq500 sequencing platform. Reads from each sample were processed using 10x Genomics Cell Ranger software (version 5.0.0). ‘cellranger mkfastq’ was used to demultiplex the Illumina sequencer’s BCL files into FASTQ files. Next, ‘cellranger count’ was used to generate single-cell gene-count matrices against the 10x Genomics pre-built GRCh38 reference genome and transcriptome (2020-A (July 7, 2020) version).

### Single cell sequencing data analysis

All data analysis and visualisation was performed using the R programming language versions 4.3.2 and 4.3.3. All of the code, figures and outputs for the analyses described herein can be viewed at the following workflowr (1.7.1) ^60^ analysis website on GitHub: https://oshlacklab.com/paediatric-cf-inflammation-citeseq/index.html. The code, as well as all necessary inputs and outputs can be cloned from the GitHub repository associated with the analysis website: https://github.com/Oshlack/paediatric-cf-inflammation-citeseq with additional data tables available at https://github.com/Oshlack/paediatric-cf-inflammation-citeseq/tree/main/output/dge_analysis. The exact R version and all package versions corresponding to each part of the analysis are specified in the “Session Info” section of each page on the analysis website.

As the data generated for this study came from two separate experiments; a small pilot (1 batch) and a larger cohort (6 batches), when discussing parts of the analysis that are unique to each experiment they will be referred to as the “scRNA-seq” (4 samples) and “CITE-seq” (41 samples) data, respectively. The analysis approaches described herein were based on the analysis we presented in Maksimovic et al., 2022 ^61^ with modifications.

#### Quality control and sample demultiplexing

The ‘DropletUtils’ (1.22.0) Bioconductor (3.18) package was used to read in the raw Cell Ranger counts and identify non-empty droplets for each individual capture. Droplets were deemed to be non-empty if they had FDR <= 0.001 as per the ‘emptyDrops’ function ^62^. 329,403 non-empty droplets were identified across all batches.

The “CITE-seq” samples were demultiplexed based on the hashtag oligo (HTO) counts using the Bioconductor ‘demuxmix’ package (1.4.0) with model=”naive”, separately for each capture ^63^. The “CITE-seq” samples were also demultiplexed using their genetic information by running ‘cellsnp-litè (1.2.0) ^64^ and ‘vireò (0.5.6) ^65^ . Specifically, reads from non-empty droplets were genotyped at 36.6M SNPs with minor allele frequency (MAF) > 0.0005 in the 1000 Genomes Project using ‘cellsnp-litè (genome1K.phase3.SNP_AF5e4.chr1toX.hg38.vcf.gz was downloaded from https://sourceforge.net/projects/cellsnp/files/SNPlist/) and then ‘vireò was used to assign each cell barcode to 1 of 8 donors, doublets, or unassigned based on these genotypes. This was run separately for each capture, and then pairs of donors were matched across captures by identifying the best match between captures based on the genotype profile of each donor.

Quality control was performed on each sample independently by visually examining the total cell number, the total unique molecular identifier (UMI) count distributions, the number of unique genes detected, and the proportions of ribosomal and mitochondrial gene counts per cell. Considering only droplets genetically assigned to a donor, outlier droplets in terms of mitochondrial read % were detected based on being more than 3 median absolute deviations (MADs) from the median value of the metric across all droplets, in each sample. Droplets that were identified as mitochondrial read % outliers were then removed, as were droplets that could not be assigned to a sample based on genetics.

To detect within-sample doublets that could not be identified from the genetic and HTO demultiplexing data, both ‘scds’ (1.18.0) ^66^ and ‘scDblFinder’ (1.16.0) ￼^67^￼^68^identified as doublets by either ‘vireò or ‘demuxmix’ (for longitudinal samples only) were filtered out. Cells that were classified as doublets by *both* ‘scds’ and ‘scDblFinder’ were also filtered out.

As significantly more cells were confidently assigned to donors using genetic demultiplexing, the genetic assignments were used in the downstream analysis for most cells; HTO assignments were used for cells from longitudinal samples. As the vast majority of genetic assignments corresponded to a single HTO, these were used to match each cell’s genetic assignment to the sample information.

Ambient RNA contamination was removed using ‘decontX’ (1.0.0) ^69^ including the raw Cell Ranger counts as background.

### Clustering and cell type annotation

The 194,407 remaining droplets were then annotated with cell type labels from the Human Lung Cell Atlas v2 (HLCA) using the ‘RunAzimuth’ function from the ‘azimuth’ (0.4.6) GitHub package. The annotated cells were divided into 3 separate data subsets based on their HLCA level 3 labels: 165,209 macrophage cells, 15,511 T/NK cells and the remaining 29,827 “rare” cells. The “rare” cells contained many cells with less than 250 reads after ambient RNA contamination, which were excluded from subsequent analysis, leaving 13,687 cells.

All data normalisation, integration and clustering was performed using functions from the ‘Seurat’ (4.4.0) CRAN package ^70–72^. For each data subset, the cell cycle scores were assigned to each cell using the ‘CellCycleScoring’ function using the inbuilt G2/M and S phase markers. Based on the “Alternate Workflow” in the ‘Seurat’ “Cell-Cycle Scoring and Regression” vignette (https://satijalab.org/seurat/articles/cell_cycle_vignette.html), the difference between the G2/M and S phase scores was calculated to maintain separation of non-cycling cells and cycling cells. The data subsets were then independently normalised using the ‘Seurat’ ‘SCTransform’ (0.4.1) function ^73,74^, regressing out the G2/M and S phase difference.

The RNA data for each cell type group was then integrated using reciprocal principal components analysis (RPCA) with 30 dimensions and 3000 features. For the T/NK and “rare” cells data, 20 neighbours were used for anchor selection (k.anchor) The number of neighbours to consider when weighting anchors (k.weight) was set to the minimum of 100 and the smallest number of cells in a single sample minus 5. For the macrophage data, default values were used for these parameters.

For the ADT data from the T/NK and “rare” subsets (excluding cells from the “scRNA-seq” experiment), all 163 ADTs were designated as variable features and the data was normalised using the centred log ratio (CLR) normalisation with margin=2. The data was then integrated using canonical correlation analysis (CCA) with 30 dimensions. Any HLA, immunoglobulin, ribosomal and mitochondrial genes were then removed from the variable features, followed by dimensionality reduction and clustering using weighted-nearest neighbor (WNN) analysis incorporating both the RNA and ADT data using the smart local moving (SLM) algorithm, with up to 30 principal components for RNA and 10 for ADT. Ten resolutions between 0.1 and 1 were explored using the ‘clustreè CRAN package (0.5.1) ^75^. A resolution of 0.6 was used in downstream analysis for T/NK and “rare” cells. This integrated and clustered data was then used as a reference that the previously excluded “scRNA-seq” cells were mapped to using the ‘MapQuery’ function from ‘Seurat’, allowing for transfer of cluster labels. A combined Uniform Manifold Approximation and Projection (UMAP) was then computed.

The macrophage data was also clustered using the SLM algorithm in ‘Seurat’, however, only the RNA data was used for clustering due to a lack of sufficiently informative ADTs for distinguishing macrophage subtypes. As previously described, a series of resolutions was examined and 0.6 was selected. The quality of the clusters in each data subset was assessed by examining their Azimuth prediction score distributions, total UMI count distributions and the distributions of the total number of unique genes detected.

Marker gene analysis was performed for each cluster, in each data subset, as described in Sim et al 2021 ^76^; log fold change cutoffs of 0.25, 0.25 and 0.5 were used in the ‘limmà(3.58.1) ^77^, ‘treat’ ^78^ test for T/NK, “rare” and macrophage cells, respectively. Gene set enrichment analysis of the REACTOME gene sets (https://www.gsea-msigdb.org/gsea/msigdb/collections.jsp) was performed for each cluster in each data subset using ‘cameraPR’ ^79^ from the ‘limmà Bioconductor package.

The ADT data from the “CITE-seq” experiment was normalised using the ‘dsb’ method (1.0.3) ^80^, including isotype controls. The top ADT markers for each cluster were then identified as described above with a log fold change cutoff of 0.1 used in the ‘treat’ test. Manual inspection of the top marker genes, ADTs and REACTOME pathways for each cluster was used to assign them cellular and/or functional identities. Clusters that shared the same manually assigned label were merged. The marker genes for the finalised clusters were identified using the ‘FindAllMarkers’ ‘Seurat’ function.

#### Differential expression analysis

Pseudobulk samples were created by summing the counts of cell type for each sample using the ‘aggregateAcrossCells’ function from the ‘scuttlè(1.12.0)^81^ Bioconductor package. Pseudobulk samples with fewer than 500 cells for total macrophages, 50 cells for macrophage subtypes and 25 cells for T/NK and “rare” groups were excluded. Only cell types with sufficient samples remaining were analysed for differential gene expression between disease conditions.

Any genes with zero counts across all pseudobulk samples were filtered out, as were genes without Entrez identifiers and those with low median log_2_ counts per million (CPM). To estimate the unwanted variation present in the data, factor analysis of “negative control features” was performed using the ‘RUVg’ function from the ‘RUVseq’(1.36.0) ^48^ Bioconductor package. The set of 3,804 human housekeeping genes published by ^82^ and provided in the ‘scMergè(1.18.0) ^83^. Bioconductor package were used as “negative control features”. Following dispersion estimation, generalized log-linear models were fitted to the counts for each gene using ‘edgeR’(4.0.15) ^49^, taking into account longitudinal samples and *k* components of unwanted variation estimated by ‘RUVg’, as well as the log_2_ of the age and the sex of each sample. The value of *k* for each analysis within a cell type was empirically determined by assessing a combination of relative log expression (RLE) and PCA plots, and p-value histograms. The Non-CF control samples were excluded from the data for the analysis of the effects of CF modifier therapy and bronchiectasis, and an additional factor for the presence of the most clinically significant microorganisms was included. Differentially expressed genes (DEG) were identified using likelihood ratio tests. As the full list of differentially expressed genes was too long to present in its entirety in Figure 3, a ‘treat’ test ^78^ with lfc=log_2_(1.1) was also applied. The Benjamini-Hochberg method ^84^ was used to adjust for multiple testing and genes with a false discovery rate (FDR) < 0.05 were deemed statistically significant.

#### Gene set and functional pathway analysis

The ‘goanà function from the ‘limmà Bioconductor package was used to test statistically significant DEGs for over-representation of gene sets. Gene sets of interest included GO, HALLMARK, REACTOME and WIKIPATHWAYS collections (v2024.1) from the Broad Institute Molecular Signatures Database ^85–87^. A list of gene sets specifically relevant to pulmonary fibrosis was also compiled and included the “defective CFTR causes cystic fibrosis” set from REACTOME, the “lung fibrosis” and “nitric oxide metabolism in cystic fibrosis” sets from WIKIPATHWAYS, a “fibrosis transcriptional signature” published by Reyfman et al 2019 ^88^ and “pulmonary fibrosis (CTD)”, which is the set of genes with direct evidence relating to pulmonary fibrosis from the Comparative Toxicogenomics Database ^89^. Multiple testing adjustment was performed using the Benjamini-Hochberg method ^84^. All of the gene sets described were also tested using a functional class scoring approach with the ‘cameraPR’ function from the ‘limmà Bioconductor package. Gene sets with an FDR < 0.05 were considered statistically significant.

#### Statistical analysis of cell type proportions

Cell type proportions were calculated using the ‘specklè (1.2.0) function ‘propeller’ ^90^, based on our broad (ann_level_1) and fine (ann_level_3) annotations, and were arcsine transformed. The transformed proportions were analysed for associations with disease conditions using the ‘limmà Bioconductor package. Briefly, linear models were fitted to the data, taking into account batch, log_2_ of the age, and sex for each sample. A random effect was also included to account for longitudinal samples using the ‘duplicateCorrelation’ function from ‘limmà. Empirical Bayes moderated t-tests with robust=TRUE ^91^ were used to identify significant changes in cell type proportion with disease condition. As above, the Benjamini-Hochberg method was used to adjust for false discovery rate ^84^. Cell types with FDR < 0.05 were considered statistically significant.

#### Statistical analysis of epithelial cells

Cell counts were log[-transformed with a pseudocount of 0.5 scaled to library size. Differential expression analysis was performed using the ‘limmà R package (3.58.1), with donor included as a covariate. Moderated t-tests were applied, incorporating the mean– variance trend and robust empirical Bayes shrinkage of variances. Significance was assessed using the TREAT method with a fold-change threshold of 1.2 and an FDR cutoff of 0.05.

## Author contributions

JM, SS, SCR, AO and MRN designed the study. SS and SCR recruited the patients and collected samples. SS, CA, DVB, and MRN performed experiments and acquired the data. JM, SS, GH, GD, PFH, AS, AO and MRN analysed the data. JM, SS, DA-Z, JEP, SCR, AO and MRN interpreted the data. All authors edited, revised, and approved the final manuscript.

## Data Availability

All data produced in the present work are contained in the manuscript, available on the analysis website provided, and on CELL x GENE.

https://oshlacklab.com/paediatric-cf-inflammation-citeseq/index.html

https://cellxgene.cziscience.com/collections/30704a65-1b50-409b-9cf6-1f26bf644c99

## Acknowledgements

The authors thank the children and parents who participated in the AREST-CF study. The authors thank the CWCCG cellular genomics team and the WEHI SCORE team.

## REFERENCES

1. Grasemann, H. & Ratjen, F. Cystic Fibrosis. The New England journal of medicine 389, 1693–1707 (2023).

2. Okuda, K., et al. Secretory Cells Dominate Airway CFTR Expression and Function in Human Airway Superficial Epithelia. American journal of respiratory and critical care medicine 203, 1275–1289 (2021).

3. Rosenow, T., et al. The cumulative effect of inflammation and infection on structural lung disease in early cystic fibrosis. The European respiratory journal 54(2019).

4. Carraro, G., et al. Transcriptional analysis of cystic fibrosis airways at single-cell resolution reveals altered epithelial cell states and composition. Nat Med 27, 806–814 (2021).

5. Schupp, J.C., et al. Single-Cell Transcriptional Archetypes of Airway Inflammation in Cystic Fibrosis. American journal of respiratory and critical care medicine 202, 1419–1429 (2020).

6. Berg, M., et al. Evidence for altered immune-structural cell crosstalk in cystic fibrosis revealed by single cell transcriptomics. Journal of cystic fibrosis: official journal of the European Cystic Fibrosis Society (2025).

7. Loske, J., et al. Pharmacological Improvement of Cystic Fibrosis Transmembrane Conductance Regulator Function Rescues Airway Epithelial Homeostasis and Host Defense in Children with Cystic Fibrosis. American journal of respiratory and critical care medicine 209, 1338–1350 (2024).

8. Januska, M.N. & Walsh, M.J. Single-Cell RNA Sequencing Reveals New Basic and Translational Insights in the Cystic Fibrosis Lung. American journal of respiratory cell and molecular biology 68, 131–139 (2023).

9. Shanthikumar, S., Stick, S.M. & Ranganathan, S.C. Minimal structural lung disease in early life represents significant pathology. Journal of Cystic Fibrosis 20, e118–e120 (2021).

10. Mall, M.A., et al. Neutrophil serine proteases in cystic fibrosis: role in disease pathogenesis and rationale as a therapeutic target. Eur Respir Rev 33(2024).

11. Malainou, C., Abdin, S.M., Lachmann, N., Matt, U. & Herold, S. Alveolar macrophages in tissue homeostasis, inflammation, and infection: evolving concepts of therapeutic targeting. J Clin Invest 133(2023).

12. Hussell, T. & Bell, T.J. Alveolar macrophages: plasticity in a tissue-specific context. Nature Reviews Immunology 14, 81–93 (2014).

13. Mould, K.J., et al. Airspace Macrophages and Monocytes Exist in Transcriptionally Distinct Subsets in Healthy Adults. American journal of respiratory and critical care medicine 203, 946–956 (2021).

14. Liao, M., et al. Single-cell landscape of bronchoalveolar immune cells in patients with COVID-19. Nature Medicine 26, 842–844 (2020).

15. Liégeois, M., et al. Airway Macrophages Encompass Transcriptionally and Functionally Distinct Subsets Altered by Smoking. American journal of respiratory cell and molecular biology 67, 241–252 (2022).

16. Liao, S.Y., et al. Single-cell RNA sequencing identifies macrophage transcriptional heterogeneities in granulomatous diseases. The European respiratory journal 57(2021).

17. Bruscia, E.M. & Bonfield, T.L. Cystic Fibrosis Lung Immunity: The Role of the Macrophage. J Innate Immun 8, 550–563 (2016).

18. Turton, K.B., Ingram, R.J. & Valvano, M.A. Macrophage dysfunction in cystic fibrosis: Nature or nurture? J Leukoc Biol 109, 573–582 (2021).

19. Squair, J.W., et al. Confronting false discoveries in single-cell differential expression. Nat Commun 12, 5692 (2021).

20. Madissoon, E., et al. A spatially resolved atlas of the human lung characterizes a gland-associated immune niche. Nat Genet 55, 66–77 (2023).

21. Sikkema, L., et al. An integrated cell atlas of the lung in health and disease. Nature Medicine 29, 1563–1577 (2023).

22. Bissonnette, E.Y., Lauzon-Joset, J.F., Debley, J.S. & Ziegler, S.F. Cross-Talk Between Alveolar Macrophages and Lung Epithelial Cells is Essential to Maintain Lung Homeostasis. Frontiers in immunology 11, 583042 (2020).

23. Morrell, E.D., et al. Cytometry TOF identifies alveolar macrophage subtypes in acute respiratory distress syndrome. JCI Insight 3(2018).

24. Li, X., et al. ScRNA-seq expression of IFI27 and APOC2 identifies four alveolar macrophage superclusters in healthy BALF. Life Sci Alliance 5(2022).

25. Oakley, M.S., et al. TCRβ-expressing macrophages induced by a pathogenic murine malaria correlate with parasite burden and enhanced phagocytic activity. PLoS One 13, e0201043 (2018).

26. Rodriguez-Cruz, A., et al. CD3(+) Macrophages Deliver Proinflammatory Cytokines by a CD3- and Transmembrane TNF-Dependent Pathway and Are Increased at the BCG-Infection Site. Frontiers in immunology 10, 2550 (2019).

27. Burel, J.G., et al. Circulating T cell-monocyte complexes are markers of immune perturbations. Elife 8(2019).

28. Yang, M., et al. Cytokine storm promoting T cell exhaustion in severe COVID-19 revealed by single cell sequencing data analysis. Precis Clin Med 5, pbac014 (2022).

29. Zheng, M.Z.M. & Wakim, L.M. Tissue resident memory T cells in the respiratory tract. Mucosal Immunol 15, 379–388 (2022).

30. Wing, E., et al. Double-negative-2 B cells are the major synovial plasma cell precursor in rheumatoid arthritis. Frontiers in immunology 14, 1241474 (2023).

31. Shanthikumar, S., et al. Inflammation in preschool cystic fibrosis is of mixed phenotype, extends beyond the lung and is differentially modified by CFTR modulators. Thorax (2025).

32. Hutt, D.M., Loguercio, S., Roth, D.M., Su, A.I. & Balch, W.E. Correcting the F508del-CFTR variant by modulating eukaryotic translation initiation factor 3-mediated translation initiation. J Biol Chem 293, 13477–13495 (2018).

33. Farinha, C.M. & Canato, S. From the endoplasmic reticulum to the plasma membrane: mechanisms of CFTR folding and trafficking. Cell Mol Life Sci 74, 39–55 (2017).

34. Kim, S.J. & Skach, W.R. Mechanisms of CFTR Folding at the Endoplasmic Reticulum. Front Pharmacol 3, 201 (2012).

35. Pardo, A., Cabrera, S., Maldonado, M. & Selman, M. Role of matrix metalloproteinases in the pathogenesis of idiopathic pulmonary fibrosis. Respiratory research 17, 23 (2016).

36. Chen, H.M., et al. Blocking immunoinhibitory receptor LILRB2 reprograms tumor-associated myeloid cells and promotes antitumor immunity. J Clin Invest 128, 5647–5662 (2018).

37. Venkataraman, T., Coleman, C.M. & Frieman, M.B. Overactive Epidermal Growth Factor Receptor Signaling Leads to Increased Fibrosis after Severe Acute Respiratory Syndrome Coronavirus Infection. J Virol 91(2017).

38. Chen, Y., et al. The potential role of CMC1 as an immunometabolic checkpoint in T cell immunity. Oncoimmunology 13, 2344905 (2024).

39. Ogilvie, V., et al. Differential global gene expression in cystic fibrosis nasal and bronchial epithelium. Genomics 98, 327–336 (2011).

40. Barros, P., Matos, A.M., Matos, P. & Jordan, P. YES1 Kinase Mediates the Membrane Removal of Rescued F508del-CFTR in Airway Cells by Promoting MAPK Pathway Activation via SHC1. Biomolecules 13(2023).

41. Paulissen, G., et al. Expression of ADAMs and their inhibitors in sputum from patients with asthma. Mol Med 12, 171–179 (2006).

42. Cote-Sierra, J., et al. Interleukin 2 plays a central role in Th2 differentiation. Proc Natl Acad Sci U S A 101, 3880–3885 (2004).

43. Zhu, J., Cote-Sierra, J., Guo, L. & Paul, W.E. Stat5 activation plays a critical role in Th2 differentiation. Immunity 19, 739–748 (2003).

44. Sun, G. & Zhou, Y.H. Identifying novel therapeutic targets in cystic fibrosis through advanced single-cell transcriptomics analysis. Comput Biol Med 187, 109748 (2025).

45. Smith, J.W., Colombo, J.L. & McDonald, T.L. Comparison of serum amyloid A and C-reactive protein as indicators of lung inflammation in corticosteroid treated and non-corticosteroid treated cystic fibrosis patients. J Clin Lab Anal 6, 219–224 (1992).

46. Hou, X., et al. CK19 stabilizes CFTR at the cell surface by limiting its endocytic pathway degradation. Faseb J 33, 12602–12615 (2019).

47. Crowell, H.L., et al. muscat detects subpopulation-specific state transitions from multi-sample multi-condition single-cell transcriptomics data. Nat Commun 11, 6077 (2020).

48. Risso, D., Ngai, J., Speed, T.P. & Dudoit, S. Normalization of RNA-seq data using factor analysis of control genes or samples. Nat Biotechnol 32, 896–902 (2014).

49. Robinson, M.D., McCarthy, D.J. & Smyth, G.K. edgeR: a Bioconductor package for differential expression analysis of digital gene expression data. Bioinformatics 26, 139–140 (2010).

50. Shanthikumar, S., Burton, M., Saffery, R., Ranganathan, S.C. & Neeland, M.R. Single-Cell Flow Cytometry Profiling of BAL in Children. American journal of respiratory cell and molecular biology 63, 152–159 (2020).

51. Jaudas, F., et al. Perinatal dysfunction of innate immunity in cystic fibrosis. Science translational medicine 17, eadk9145 (2025).

52. Gaggar, A., et al. The role of matrix metalloproteinases in cystic fibrosis lung disease. European Respiratory Journal 38, 721–727 (2011).

53. Bühling, F., et al. Pivotal role of cathepsin K in lung fibrosis. Am J Pathol 164, 2203–2216 (2004).

54. Shanthikumar, S., Ranganathan, S. & Neeland, M.R. Ivacaftor, not ivacaftor/lumacaftor, associated with lower pulmonary inflammation in preschool cystic fibrosis. Pediatric pulmonology 57, 2549–2552 (2022).

55. Shanthikumar, S. & Massie, J. A Review of Treatments That Improve Cystic Fibrosis Transmembrane Conductance Regulator Function. Clinical Medicine Insights: Therapeutics 9, 1179559X17719123 (2017).

56. Zhang, S., et al. Cystic fibrosis macrophage function and clinical outcomes after elexacaftor/tezacaftor/ivacaftor. The European respiratory journal 61(2023).

57. Quon, B.S. & Rowe, S.M. New and emerging targeted therapies for cystic fibrosis. Bmj 352, i859 (2016).

58. Sly, P.D., et al. Risk factors for bronchiectasis in children with cystic fibrosis. The New England journal of medicine 368, 1963–1970 (2013).

59. Shivanthan Shanthikumar, L.G., Melanie R Neeland Processing of pediatric bronchoalveolar lavage samples for single cell analysis v3. *protocols.io* (2024).

60. Blischak, J.D., Carbonetto, P. & Stephens, M. Creating and sharing reproducible research code the workflowr way. F1000Research 8, 1749 (2019).

61. Maksimovic, J., et al. Single-cell atlas of bronchoalveolar lavage from preschool cystic fibrosis reveals new cell phenotypes. bioRxiv, 2022.2006.2017.496207 (2022).

62. Lun, A.T.L., et al. EmptyDrops: distinguishing cells from empty droplets in droplet-based single-cell RNA sequencing data. Genome Biol 20, 63 (2019).

63. Klein, H.U. demuxmix: Demultiplexing oligonucleotide-barcoded single-cell RNA sequencing data with regression mixture models. bioRxiv (2023).

64. Huang, X. & Huang, Y. Cellsnp-lite: an efficient tool for genotyping single cells. Bioinformatics 37, 4569–4571 (2021).

65. Huang, Y., McCarthy, D.J. & Stegle, O. Vireo: Bayesian demultiplexing of pooled single-cell RNA-seq data without genotype reference. Genome Biol 20, 273 (2019).

66. Bais, A.S. & Kostka, D. scds: computational annotation of doublets in single-cell RNA sequencing data. Bioinformatics 36, 1150–1158 (2020).

67. Germain, P.L., Lun, A., Garcia Meixide, C., Macnair, W. & Robinson, M.D. Doublet identification in single-cell sequencing data using scDblFinder. F1000Research 10, 979 (2021).

68. Neavin, D., et al. Demuxafy: improvement in droplet assignment by integrating multiple single-cell demultiplexing and doublet detection methods. Genome Biol 25, 94 (2024).

69. Yang, S., et al. Decontamination of ambient RNA in single-cell RNA-seq with DecontX. Genome Biol 21, 57 (2020).

70. Hao, Y., et al. Integrated analysis of multimodal single-cell data. 2020.2010.2012.335331 (2020).

71. Butler, A., Hoffman, P., Smibert, P., Papalexi, E. & Satija, R. Integrating single-cell transcriptomic data across different conditions, technologies, and species. Nat Biotechnol 36, 411–420 (2018).

72. Stuart, T., et al. Comprehensive Integration of Single-Cell Data. Cell 177, 1888–1902.e1821 (2019).

73. Choudhary, S. & Satija, R. Comparison and evaluation of statistical error models for scRNA-seq. Genome Biol 23, 27 (2022).

74. Hafemeister, C. & Satija, R. Normalization and variance stabilization of single-cell RNA-seq data using regularized negative binomial regression. Genome Biol 20, 296 (2019).

75. Zappia, L. & Oshlack, A. Clustering trees: a visualization for evaluating clusterings at multiple resolutions. GigaScience 7(2018).

76. Sim, C.B., et al. Sex-Specific Control of Human Heart Maturation by the Progesterone Receptor. Circulation (2021).

77. Ritchie, M.E., et al. limma powers differential expression analyses for RNA-sequencing and microarray studies. Nucleic acids research 43, e47 (2015).

78. McCarthy, D.J. & Smyth, G.K. Testing significance relative to a fold-change threshold is a TREAT. Bioinformatics 25, 765–771 (2009).

79. Wu, D. & Smyth, G.K. Camera: a competitive gene set test accounting for inter-gene correlation. Nucleic acids research 40, e133 (2012).

80. Mulè, M.P., Martins, A.J. & Tsang, J.S. Normalizing and denoising protein expression data from droplet-based single cell profiling. Nat Commun 13, 2099 (2022).

81. McCarthy, D.J., Campbell, K.R., Lun, A.T. & Wills, Q.F. Scater: pre-processing, quality control, normalization and visualization of single-cell RNA-seq data in R. Bioinformatics 33, 1179–1186 (2017).

82. Eisenberg, E. & Levanon, E.Y. Human housekeeping genes, revisited. Trends Genet 29, 569–574 (2013).

83. Lin, Y., et al. scMerge leverages factor analysis, stable expression, and pseudoreplication to merge multiple single-cell RNA-seq datasets. Proc Natl Acad Sci U S A 116, 9775–9784 (2019).

84. Benjamini, Y. & Hochberg, Y. Controlling the False Discovery Rate - a Practical and Powerful Approach to Multiple Testing. J R Stat Soc B 57, 289–300 (1995).

85. Liberzon, A., et al. The Molecular Signatures Database (MSigDB) hallmark gene set collection. Cell Syst 1, 417–425 (2015).

86. Liberzon, A., et al. Molecular signatures database (MSigDB) 3.0. Bioinformatics 27, 1739–1740 (2011).

87. Subramanian, A., et al. Gene set enrichment analysis: a knowledge-based approach for interpreting genome-wide expression profiles. Proc Natl Acad Sci U S A 102, 15545–15550 (2005).

88. Reyfman, P.A., et al. Single-Cell Transcriptomic Analysis of Human Lung Provides Insights into the Pathobiology of Pulmonary Fibrosis. American journal of respiratory and critical care medicine 199, 1517–1536 (2019).

89. Davis, A.P., et al. Comparative Toxicogenomics Database’s 20th anniversary: update 2025. Nucleic acids research 53, D1328–d1334 (2025).

90. Phipson, B., et al. propeller: testing for differences in cell type proportions in single cell data. Bioinformatics 38, 4720–4726 (2022).

91. Phipson, B., Lee, S., Majewski, I.J., Alexander, W.S. & Smyth, G.K. ROBUST HYPERPARAMETER ESTIMATION PROTECTS AGAINST HYPERVARIABLE GENES AND IMPROVES POWER TO DETECT DIFFERENTIAL EXPRESSION. Ann Appl Stat 10, 946–963 (2016).

