## Supplementary Figures for "Single-cell profiling of BAL in preschool cystic fibrosis reveals macrophage dysregulation and ivacaftor-modified inflammatory programs in the early life lung"

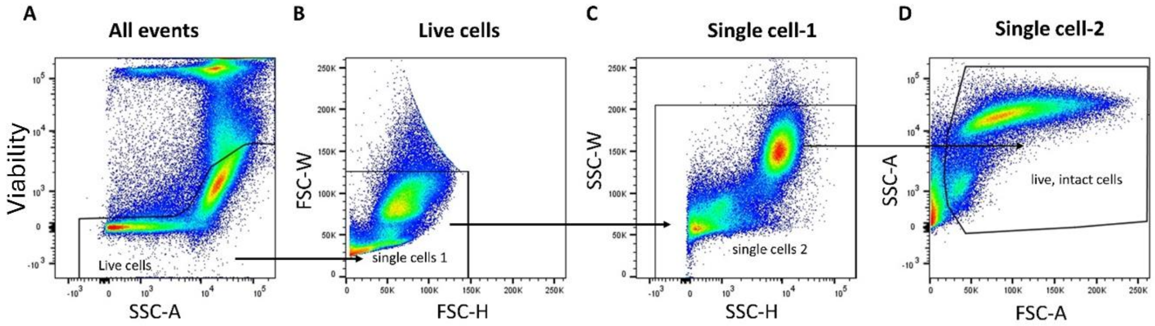


**Supplementary Figure 1.** Gating strategy used to sort live, single BAL cells for downstream single-cell sequencing. (A) Firstly, live cells were selected using SSC-A vs Viability. (B-C) Within the live cell gate, single cells were identified using FSC-H vs W, followed by SSC-H vs W. (D) Finally, debris was removed, and live, single intact cells were selected.

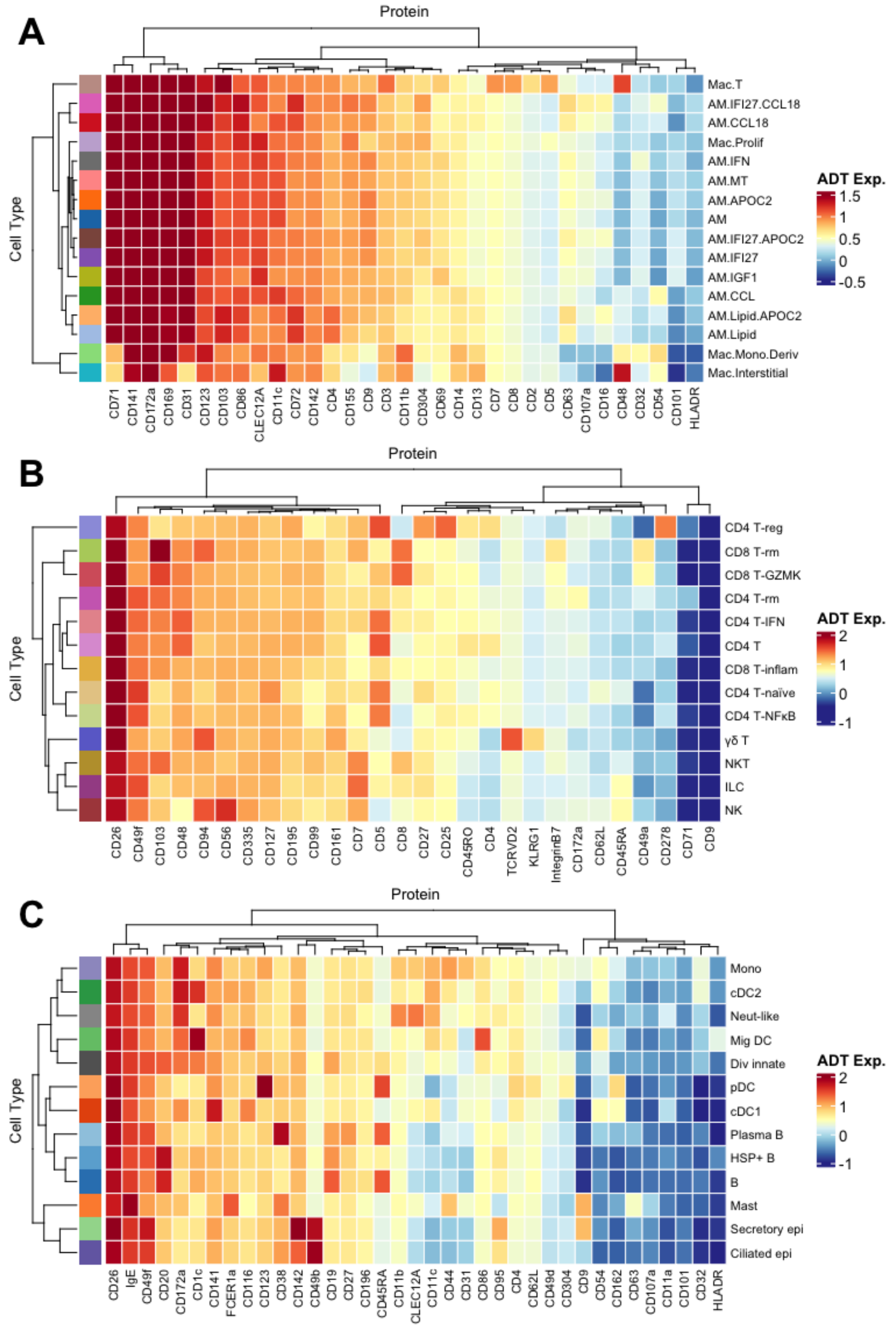


**Supplementary Figure 2**. Protein (antibody derived tag (ADT)) expression on cell populations identified during sub-clustering analysis of **(A)** BAL macrophages, **(B)** BAL T/NK cells and **(C)** myeloid/rare cells.

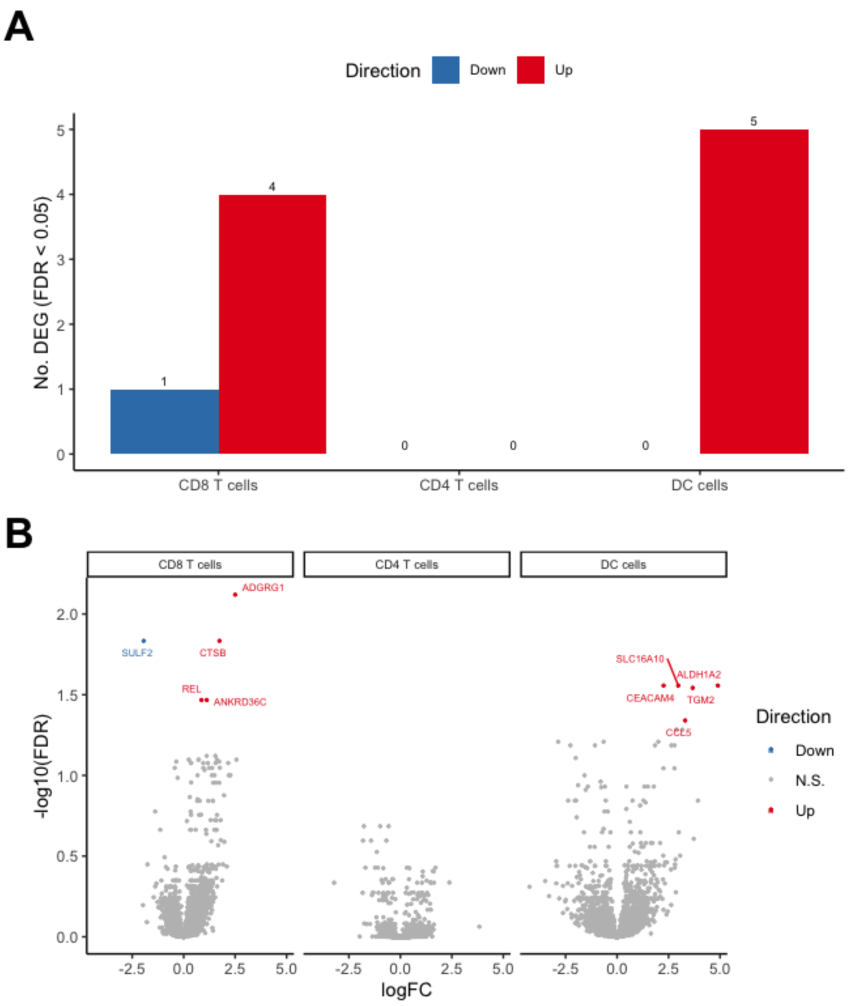


C

Differentially expressed genes by cell type

CF.NO_MODvNON_CF.CTRL

| gene | logFC | FDR |
|---|---|---|
| **CD8 T cells** | | |
| ADGRG1 | 2.5021012 | 0.00758679 |
| SULF2 | -1.9413497 | 0.01467086 |
| CTSB | 1.7399768 | 0.01467086 |
| REL | 0.8640961 | 0.03411823 |
| ANKRD36C | 1.1218820 | 0.03411823 |
| **DC cells** | | |
| CEACAM4 | 2.2610736 | 0.02775101 |
| SLC16A10 | 2.9741874 | 0.02775101 |
| ALDH1A2 | 4.8973876 | 0.02775101 |
| TGM2 | 3.6728039 | 0.02868435 |
| CCL5 | 3.3089485 | 0.04566559 |

**Supplementary Figure 3.** Differential expression analysis between CF (no mod) (n=25) and non-CF control (n=8) samples for CD8 T cells, CD4 T cells, and dendritic cells (DC cells). **(A)** Summary of the number of genes differentially expressed genes in CD8 T cells, CD4 T cells and DC cells. **(B)** Volcano plot and summary table of the details related to the differentially expressed genes.

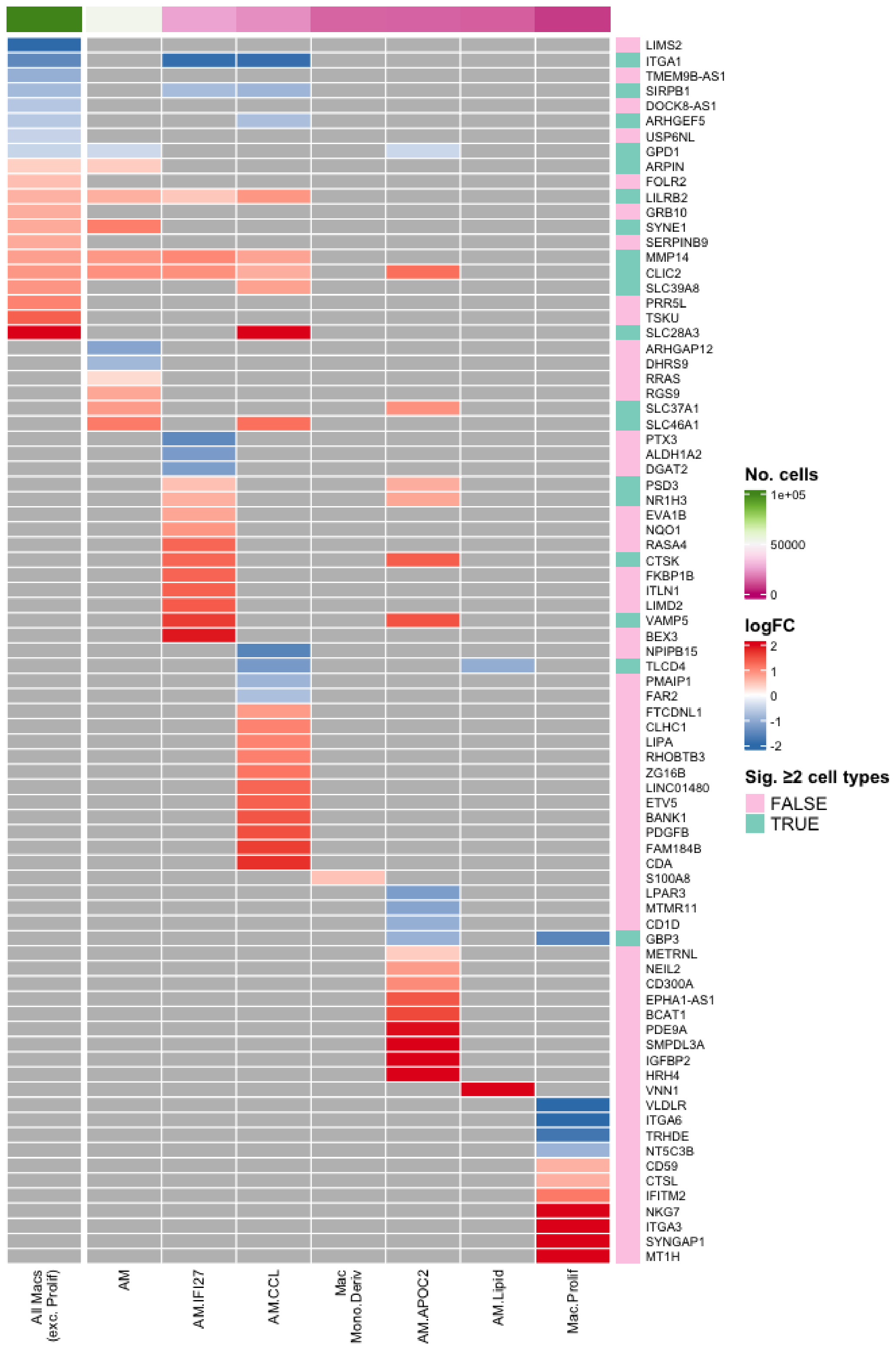


**Supplementary Figure 4.** Full list of differentially expressed genes within each tested macrophage subpopulation for the CF (no mod)(S) "severe" vs CF (no mod)(M) "mild" comparison in Figure 4.

A

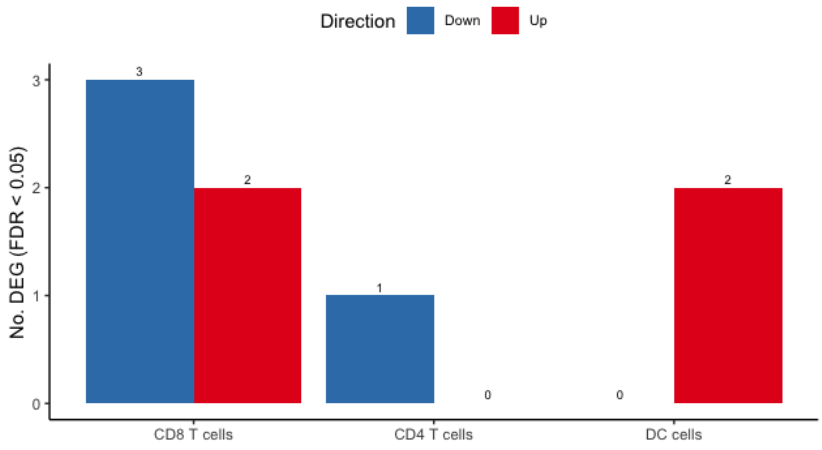


B

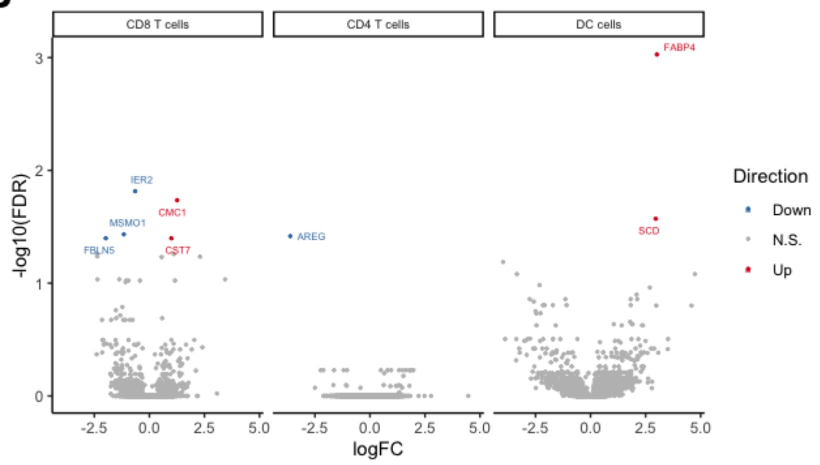


C

**Differentially expressed genes by cell type**
CF.NO_MOD.SvCF.NO_MOD.M

| gene | logFC | FDR |
|---|---|---|
| **CD8 T cells** | | |
| IER2 | -0.6465572 | 0.0152957508 |
| CMC1 | 1.2609872 | 0.0184005852 |
| MSMO1 | -1.1619299 | 0.0368874786 |
| FBLN5 | -1.9805997 | 0.0399874695 |
| CST7 | 1.0002086 | 0.0399874695 |
| **CD4 T cells** | | |
| AREG | -3.6207178 | 0.0382842050 |
| **DC cells** | | |
| FABP4 | 3.0135376 | 0.0009380894 |
| SCD | 2.9594586 | 0.0268390379 |

**Supplementary Figure 5.** Differential expression analysis between CF (no mod)(S) “severe” (n=8) and CF (no mod)(M) “mild” (n=17) samples for CD8 T cells, CD4 T cells, and dendritic cells (DC cells). **(A)** Summary of the number of genes differentially expressed genes in CD8 T cells, CD4 T cells and DC cells. **(B)** Volcano plot and summary table of the details related to the differentially expressed genes.

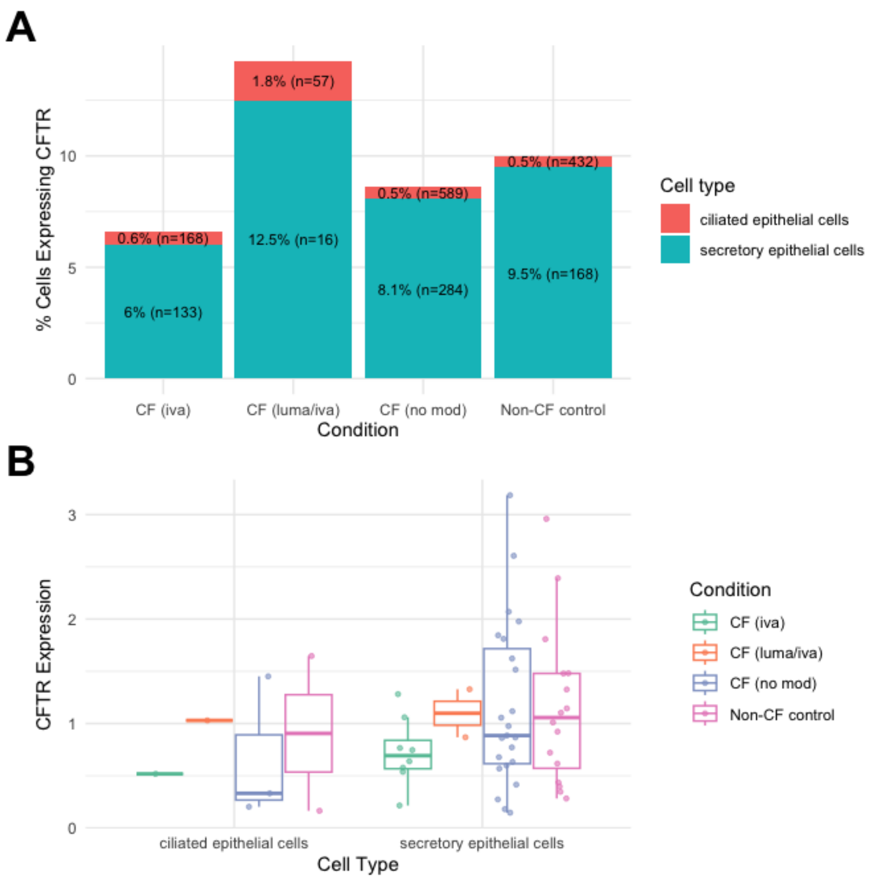


**Supplementary Figure 6.** CFTR in epithelial cells. (A) Percentage of CFTR-expressing cells for each epithelial cell subset. (B) Log CFTR expression, only for CFTR +ve cells.

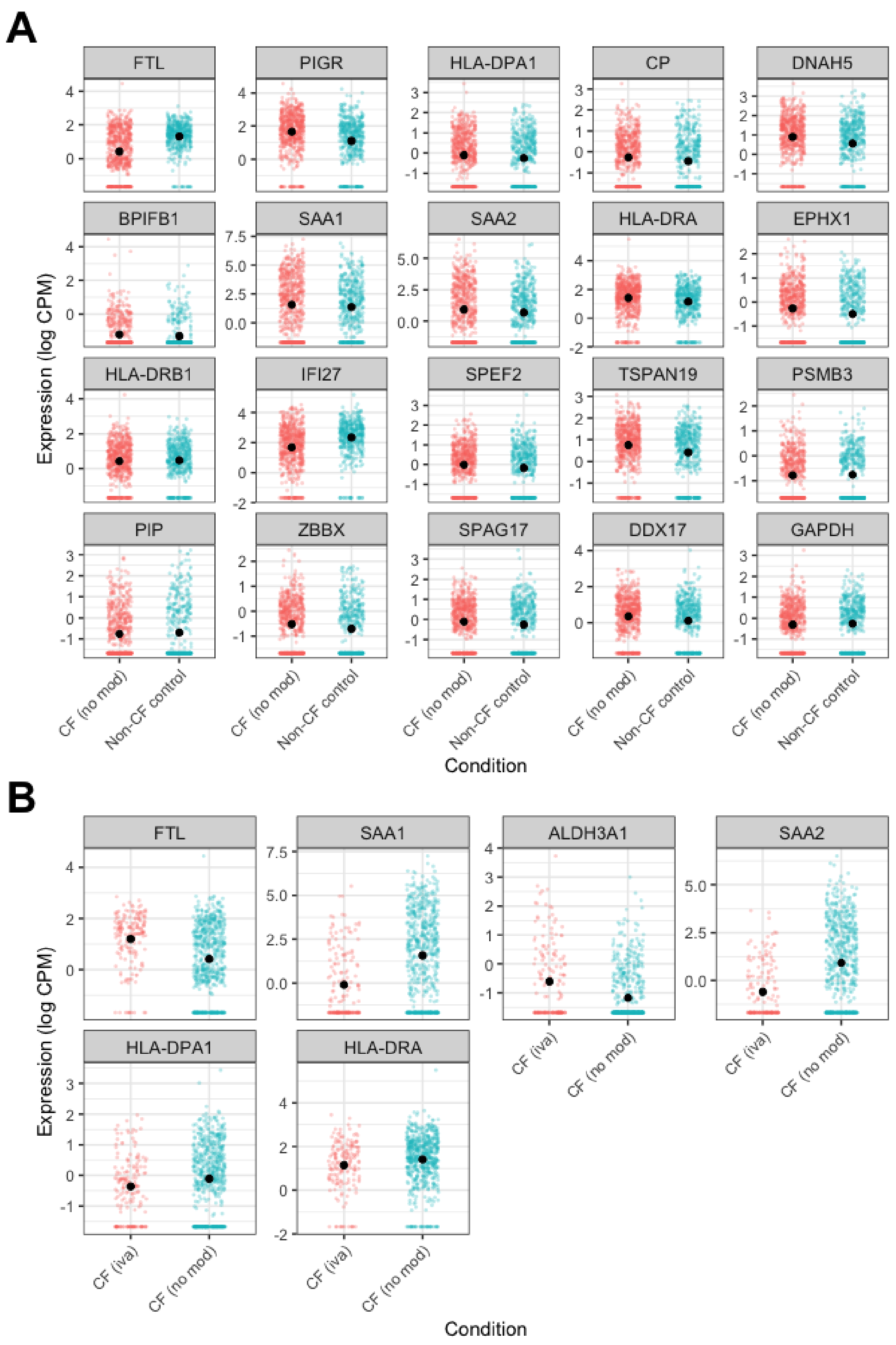


**Supplementary Figure 7.** Significantly differentially expressed genes (DEGs) from the Sun and Zhou (2025) and Carraro et al. (2021) sets in ciliated epithelial cells. DEGs were significant at FDR < 0.05 relative to a fold change of 1.2. (A) Significant DEGs for non-CF controls vs. CF (no mod). Black dot = group mean. (B) Significant DEGs for CF (iva) vs. CF (no mod). Black dot = group mean.

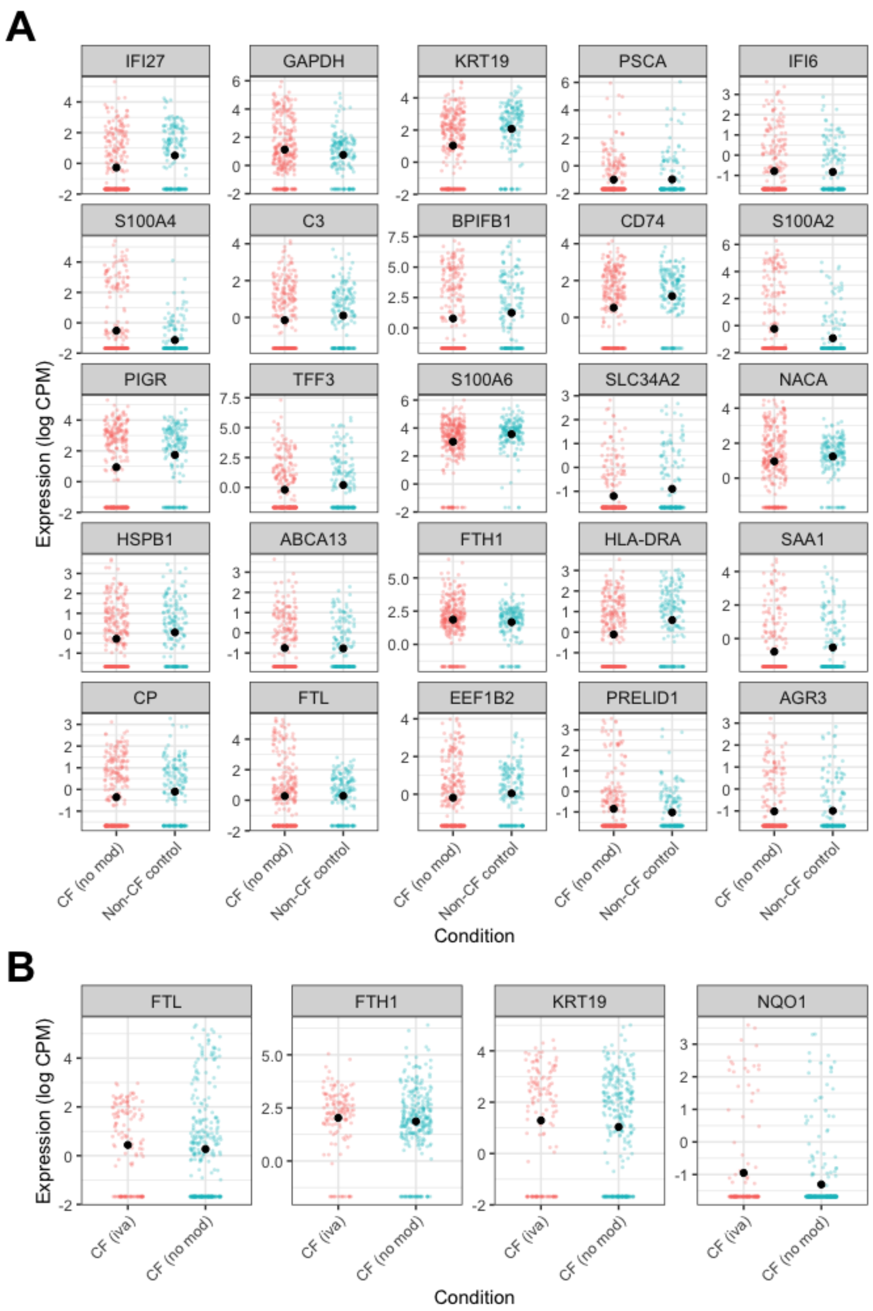


**Supplementary Figure 8.** Significantly differentially expressed genes (DEGs) from the Sun and Zhou (2025) and Carraro et al. (2021) sets in secretory epithelial cells. DEGs were significant at FDR < 0.05 relative to a fold change of 1.2. (A) Significant DEGs for non-CF controls vs. CF (no mod). Black dot = group mean. (B) Significant DEGs for CF (iva) vs. CF (no mod). Black dot = group mean.
